# Mpox diagnosis history, behavioural risk modification, and vaccination uptake in gay, bisexual, and other men who have sex with men in the UK: findings from a large, online community cross-sectional survey (RiiSH-Mpox) undertaken November/December 2022

**DOI:** 10.1101/2023.05.11.23289797

**Authors:** Dana Ogaz, Qudsia Enayat, Jack RG Brown, Dawn Phillips, Ruth Wilkie, Danielle Jayes, David Reid, Gwenda Hughes, Catherine H Mercer, John Saunders, Hamish Mohammed, UKHSA Sexual Health Liaison Group

## Abstract

Mpox cases, first identified as part of the multi-country outbreak in May 2022, rapidly fell in the UK from September 2022. Public health responses included community-coordinated messaging and a targeted mpox vaccination in eligible gay, bisexual, and other men who have sex with men (GBMSM). Using data from an online, community survey of GBMSM (November/December 2022), we examined self-reported mpox diagnoses, behavioural risk modification, and mpox vaccination offer and uptake (≥1 dose). Thirty-five participants (2.6%, 35/1,333) were ever mpox test positive; half (53%, 707/1,333) reported behaviour modification to avoid mpox. In GBMSM considered vaccine eligible, uptake was 69% (95% CI: 65%-72%, 601/875) or 92% (95% CI: 89%-94%, 601/655) in those eligible and offered. GBMSM self-identifying as bisexual, those with lower educational qualifications, and those unemployed were less likely to be vaccinated. Equitable mpox vaccine offer and provision is needed to minimise the risk of future outbreaks and mpox-related health inequalities.

## Introduction

On 23 July 2022, the World Health Organization (WHO) declared the global mpox (formerly ‘monkeypox’) outbreak a Public Health Emergency of International Concern (1). To date, there have been over 85,000 confirmed mpox cases across 110 countries (2). Mpox virus can be transmitted through close, skin-to-skin contact with an infected individual (1). The 2022 outbreak was the first widespread international outbreak of mpox predominantly transmitted through sexually associated contact and disproportionately affecting gay, bisexual and other men who have sex with men (GBMSM) (1, 3–5).

Mpox was first detected in the UK in May 2022, but community transmission has been estimated to have started during the month prior (3, 5). Case numbers peaked in July 2022 and reached over 3,500 cases across the UK through year end. The UK Health Security Agency (UKHSA) enacted measures to curb rising infection rates, including raising public health awareness, advising a self-isolation period of 21 days for people with a diagnosis, comprehensive contact tracing of recent close sexual contacts, and a recommendation of a targeted vaccination campaign utilising Modified Vaccinia Ankara vaccine (6). UKHSA worked closely with community-based organisations to inform public health messaging to raise awareness and reduce the risk of mpox (7–9). An expert consensus panel, which included UKHSA, along with national and community sexual health organisations, estimated that 111,000 people would be eligible for mpox vaccination in the UK, including 103,000 GBMSM and 8,000 healthcare and outreach workers (10). The vaccination campaign, which began in June 2022 had vaccinated 70,837 people with a first dose, and 31,827 with a second through May 2023 (11). Case numbers in the UK rapidly subsided by the end of September 2022, with few reported cases in 2023 (12).

Vaccine delivery was initially targeted to London sexual health services (SHS), as residents of the capital comprised at least 69% of cases through September 2022 (10). National guidance recommended GBMSM at highest risk of mpox could be identified among those attending SHS, using similar markers of risk utilised in identifying those eligible for HIV pre-exposure prophylaxis (PrEP), irrespective of HIV status (6, 13). Vaccine eligibility criteria included a recent history of multiple recent sexual partners, group sex, attending sex-on-premises venues or diagnosis of a bacterial sexually transmitted infection (STI). First doses were initially offered to eligible patients as a subcutaneous injection (6, 14), with fractional dosing via intradermal administration also piloted and rolled out to maximise the coverage of the available vaccine supply (15). Observational, real-world studies have since reported 78%-86% vaccine effectiveness in preventing symptomatic mpox infection (16, 17).

In response to the UK mpox outbreak, the “Reducing inequalities in Sexual Health (RiiSH)-Mpox” survey was rapidly deployed from 24 November-19 December 2022 to assess the impact of the mpox outbreak on the health and well-being, sexual behaviour, and service-use among a community sample of GBMSM in the UK. We used these survey data to examine self-reported mpox diagnosis history, behavioural risk modification, and uptake of mpox vaccination among GBMSM in the UK.

## Methods

### Data collection and study design

The RiiSH-Mpox survey was adapted from an established methodology used to deliver a series of repeat, cross-sectional RiiSH surveys carried out initially in 2017 and during periods before and after COVID-19 related social restrictions in the UK (18, 19). The RiiSH-Mpox survey included all previous questions on service use and sexual risk behaviour but incorporated a novel module, developed with input from community stakeholders, relating to mpox diagnosis, vaccination uptake and behavioural risk modification in response to the mpox outbreak.

### Setting and sampling

As in previous rounds of the RiiSH surveys, participants were recruited through adverts on social networking sites (Facebook, Instagram and Twitter) and Grindr, a geospatial networking (dating) application. Those included in analyses were aged ≥16 years, UK residents, self-identifying as men (cisgender/transgender), transgender women, or gender-diverse individuals assigned male at birth (AMAB), who reported having had sex with a man (cisgender/transgender) or gender-diverse individual AMAB in the last year. Online consent was obtained from all participants and no incentive was offered to participate.

### Data analysis

#### Mpox testing and diagnosis history

We calculated the percentage and 95% confidence interval (CI) (Clopper-Pearson) of those ever reporting a mpox diagnosis history (i.e., positive mpox test) through survey completion. As a sensitivity analysis, we calculated the percentage reporting a diagnosis history or self-perceived mpox in absence of a positive mpox test or self-reported testing history.

Using Pearson’s chi-squared test, we assessed differences in sociodemographic, clinical, and behavioural characteristics (described below). Given the low number of participants with a diagnosis history, regression analyses were not conducted.

#### Mpox vaccination uptake

Mpox vaccination uptake was defined as the receipt of ≥1 mpox vaccine doses. Report of vaccine offer (“Have you been offered a vaccine for monkeypox?”) was assessed in those not reporting uptake. We present the percentage and 95% CI of uptake among those offered a mpox vaccine and in all participants.

#### Mpox vaccination uptake in those mpox vaccination eligible

We examined uptake among those assumed to be eligible for mpox vaccination based on equivalent or proxy criteria outlined in national vaccination guidance (13). Using survey responses, vaccine eligibility was defined as the report of any of the following since August 2022: ≥10 physical male sex partners meeting any physical male sex partner(s) in a sex on premises venue, sex party, or cruising grounds (hereafter, public sex environment [PSE]); a positive STI test; or in the last year, report of: PrEP use (as a proxy for those at higher risk of mpox acquisition); or use of recreational drugs associated with chemsex (crystal methamphetamine, mephedrone or gamma-hydroxybutyrate/gamma-butyrolactone). As a sensitivity analysis, we used a lower threshold of ≥5 physical male sex partners since August 2022, instead of ≥10, to define eligibility.

#### Factors associated with mpox vaccination

We assessed factors associated with mpox vaccination using Pearson’s chi-squared test and binary logistic regression. Sociodemographic variables with a significant association with vaccination in bivariate analysis were included in multivariable regression models sequentially assessing associations of key clinical and behavioural characteristics with mpox vaccination. Age-group and ethnicity were selected *a priori* for inclusion in multivariable modelling. A sensitivity analysis, examining sociodemographic factors associated with mpox vaccination in those considered mpox eligible, was also carried out.

Sociodemographic characteristics used in descriptive and regression analyses included: age-group, ethnicity, gender, sexual orientation, country of birth, UK nation of residence (England, Scotland, Wales, or Northern Ireland), education-level, employment, household composition (living alone or not), relationship status (single or in a relationship), and report of a comfortable financial situation (top two quartiles [“I am comfortable”/“I am very comfortable”] from the question, “How would you best describe your current financial situation”). Clinical characteristics included: HIV status, uptake of ≥1 Hepatitis A vaccine dose(s), uptake of ≥1 Hepatitis B vaccine(s), and uptake of ≥1 human papillomavirus [HPV] vaccine doses (or the report of any in multivariable analyses). Behavioural risk modification as a result of the mpox outbreak was defined as the report of any of the following since May 2022: fewer sexual partners, reduced visits to sex on premises venues or PSE, and avoiding the following: any sex, condomless anal sex, skin-to-skin contact, visiting clubs or crowds. Sexual risk behaviour indicators, comprising mpox vaccination eligibility proxies, included: number and meeting place of male physical sex partners since August 2022, a positive STI test since August 2022, and in the last year, the report of PrEP use, or recreational drug use associated with chemsex. Lookback periods varied across sexual risk indicators to match those used in prior waves of the RiiSH survey or to focus on behaviours since the start of the mpox outbreak in May.

Due to small participant numbers in subgroups, we dichotomised ethnic (White, all other ethnic groups), gender (cisgender male, all other gender identity groups), and sexual orientation groups (gay/homosexual, bisexual [which includes those identifying as bisexual, straight, or “another way”]) for analyses.

Survey data were collected via the Snap Surveys platform and all data management and analyses were undertaken using Stata v15.0 (StataCorp, College Station, TX, USA). All p-values less than 5% were considered statistically significant.

## Results

Of the 1,435 GBMSM that engaged with the RiiSH-Mpox survey in December 2022, 1,333 (93%) met gender identity and sexual history selection criteria to participate (Appendix I). Missing data were limited (<3% item non-response) as most survey questions were compulsory. Participants had a median age of 45 years (interquartile range [IQR]: 35-55, range: 16-78) (Table 1,4). Most self-identified as cisgender male (99%), gay/homosexual (89%), were of White ethnicity (92%), lived in England (86%), and were employed (81%). Nearly half reported a comfortable financial situation (48%), two-thirds had degree-level qualifications (63%), and 15% of participants were living with HIV. Most participants were recruited from Facebook (58%), followed by Grindr (24%) and Twitter (15%). Of all participants, 53% (707/1,333) reported behavioural modification to avoid getting mpox, where the reduction in number of physical male sex partners (72% 510/707) was the most reported measure (Table 2).

**Table 1.**
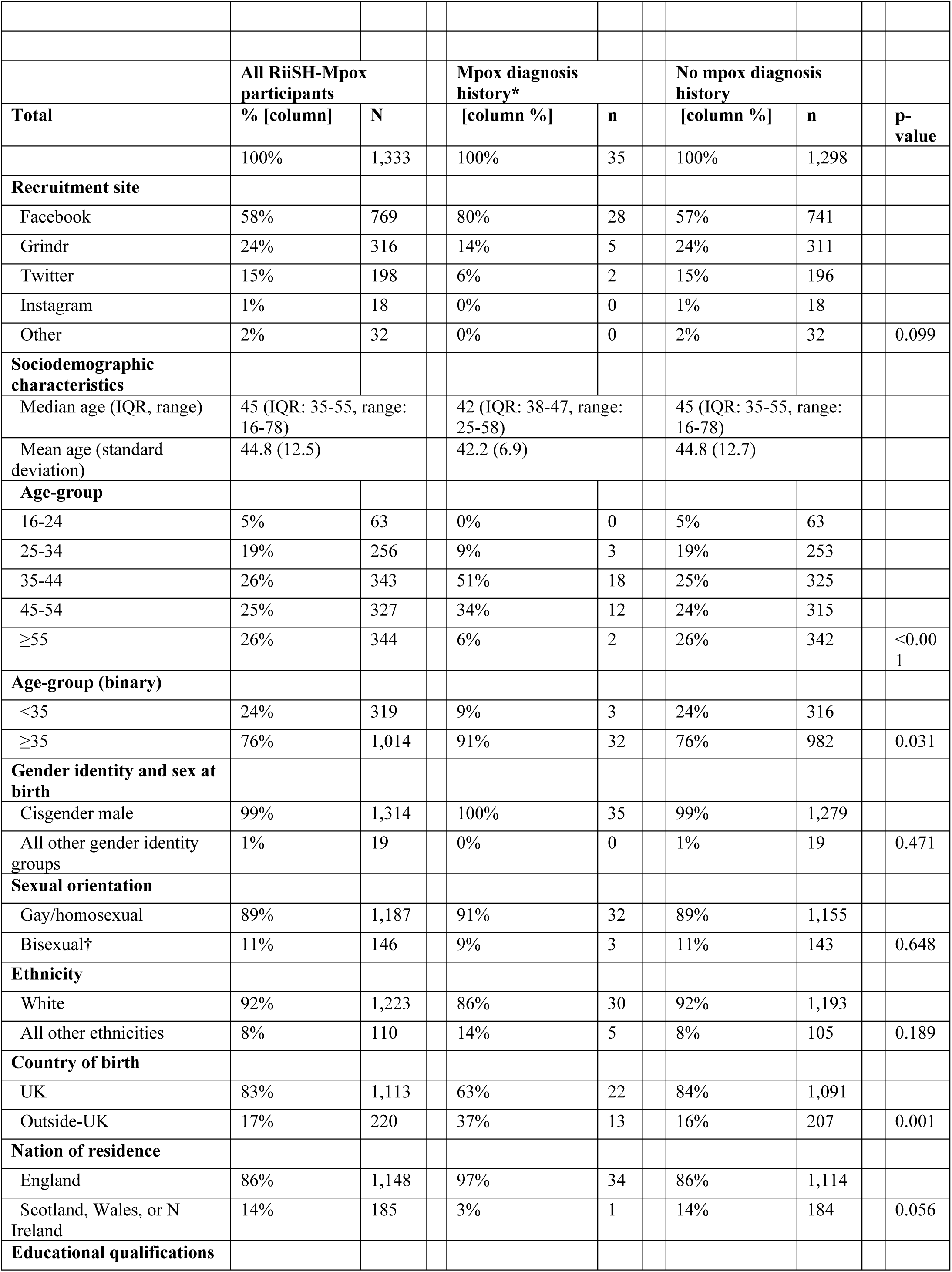
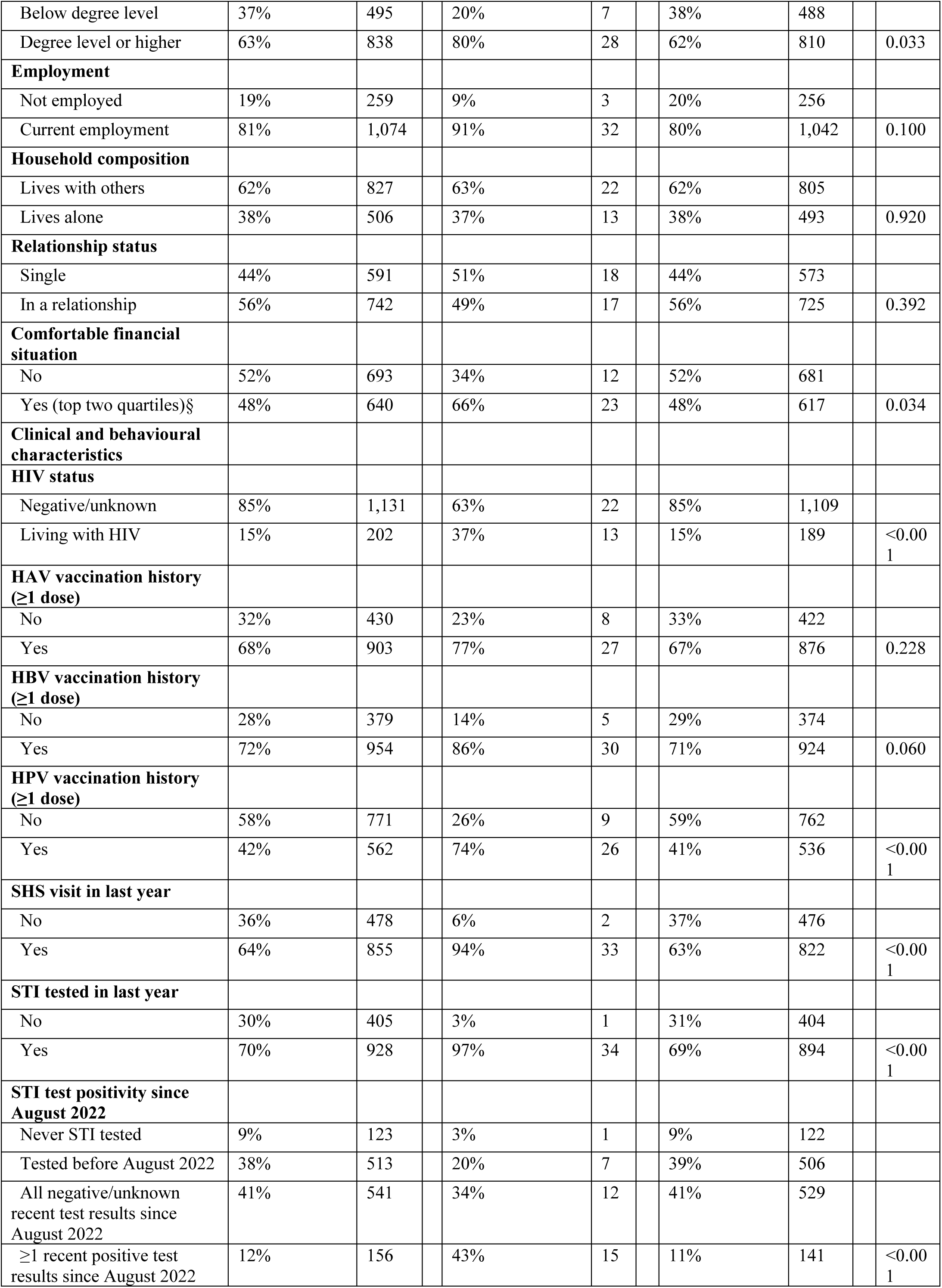
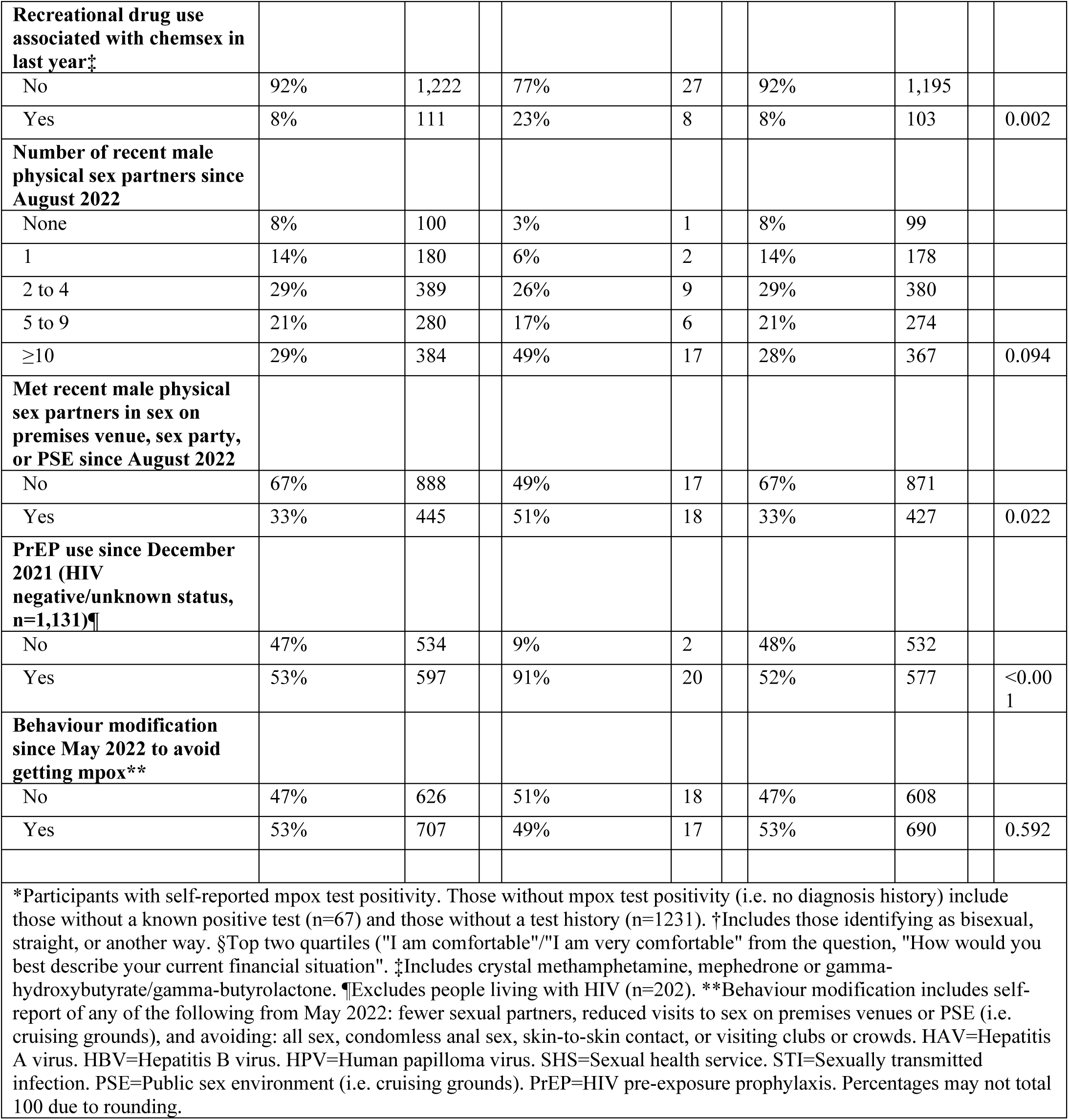
Sociodemographic and clinical characteristics of RiiSH-Mpox participants with and without a mpox diagnosis history, November/December 2022.

|  |  |  |  |  |  |  |  |  |
| --- | --- | --- | --- | --- | --- | --- | --- | --- |
|  | **All RiiSH-Mpox participants** |  | **Mpox diagnosis history\*** |  | **No mpox diagnosis history** |  |  |  |
| **Total** | **% [column]** | **N** | **[column %]** | **n** | **[column %]** | **n** |  | **p-value** |
|  | 100% | 1,333 | 100% | 35 | 100% | 1,298 |  |  |
| **Recruitment site** |  |  |  |  |  |  |  |  |
| Facebook | 58% | 769 | 80% | 28 | 57% | 741 |  |  |
| Grindr | 24% | 316 | 14% | 5 | 24% | 311 |  |  |
| Twitter | 15% | 198 | 6% | 2 | 15% | 196 |  |  |
| Instagram | 1% | 18 | 0% | 0 | 1% | 18 |  |  |
| Other | 2% | 32 | 0% | 0 | 2% | 32 |  | 0.099 |
| **Sociodemographic characteristics** |  |  |  |  |  |  |  |  |
| Median age (IQR, range) | 45 (IQR: 35-55, range: 16-78) |  | 42 (IQR: 38-47, range: 25-58) |  | 45 (IQR: 35-55, range: 16-78) |  |  |  |
| Mean age (standard deviation) | 44.8 (12.5) |  | 42.2 (6.9) |  | 44.8 (12.7) |  |  |  |
| **Age-group** |  |  |  |  |  |  |  |  |
| 16-24 | 5% | 63 | 0% | 0 | 5% | 63 |  |  |
| 25-34 | 19% | 256 | 9% | 3 | 19% | 253 |  |  |
| 35-44 | 26% | 343 | 51% | 18 | 25% | 325 |  |  |
| 45-54 | 25% | 327 | 34% | 12 | 24% | 315 |  |  |
| ≥55 | 26% | 344 | 6% | 2 | 26% | 342 |  | <0.001 |
| **Age-group (binary)** |  |  |  |  |  |  |  |  |
| <35 | 24% | 319 | 9% | 3 | 24% | 316 |  |  |
| ≥35 | 76% | 1,014 | 91% | 32 | 76% | 982 |  | 0.031 |
| **Gender identity and sex at birth** |  |  |  |  |  |  |  |  |
| Cisgender male | 99% | 1,314 | 100% | 35 | 99% | 1,279 |  |  |
| All other gender identity groups | 1% | 19 | 0% | 0 | 1% | 19 |  | 0.471 |
| **Sexual orientation** |  |  |  |  |  |  |  |  |
| Gay/homosexual | 89% | 1,187 | 91% | 32 | 89% | 1,155 |  |  |
| Bisexual† | 11% | 146 | 9% | 3 | 11% | 143 |  | 0.648 |
| **Ethnicity** |  |  |  |  |  |  |  |  |
| White | 92% | 1,223 | 86% | 30 | 92% | 1,193 |  |  |
| All other ethnicities | 8% | 110 | 14% | 5 | 8% | 105 |  | 0.189 |
| **Country of birth** |  |  |  |  |  |  |  |  |
| UK | 83% | 1,113 | 63% | 22 | 84% | 1,091 |  |  |
| Outside-UK | 17% | 220 | 37% | 13 | 16% | 207 |  | 0.001 |
| **Nation of residence** |  |  |  |  |  |  |  |  |
| England | 86% | 1,148 | 97% | 34 | 86% | 1,114 |  |  |
| Scotland, Wales, or N Ireland | 14% | 185 | 3% | 1 | 14% | 184 |  | 0.056 |
| **Educational qualifications** |  |  |  |  |  |  |  |  |
| Below degree level | 37% | 495 | 20% | 7 | 38% | 488 |  |  |
| Degree level or higher | 63% | 838 | 80% | 28 | 62% | 810 |  | 0.033 |
| **Employment** |  |  |  |  |  |  |  |  |
| Not employed | 19% | 259 | 9% | 3 | 20% | 256 |  |  |
| Current employment | 81% | 1,074 | 91% | 32 | 80% | 1,042 |  | 0.100 |
| **Household composition** |  |  |  |  |  |  |  |  |
| Lives with others | 62% | 827 | 63% | 22 | 62% | 805 |  |  |
| Lives alone | 38% | 506 | 37% | 13 | 38% | 493 |  | 0.920 |
| **Relationship status** |  |  |  |  |  |  |  |  |
| Single | 44% | 591 | 51% | 18 | 44% | 573 |  |  |
| In a relationship | 56% | 742 | 49% | 17 | 56% | 725 |  | 0.392 |
| **Comfortable financial situation** |  |  |  |  |  |  |  |  |
| No | 52% | 693 | 34% | 12 | 52% | 681 |  |  |
| Yes (top two quartiles)§ | 48% | 640 | 66% | 23 | 48% | 617 |  | 0.034 |
| **Clinical and behavioural characteristics** |  |  |  |  |  |  |  |  |
| **HIV status** |  |  |  |  |  |  |  |  |
| Negative/unknown | 85% | 1,131 | 63% | 22 | 85% | 1,109 |  |  |
| Living with HIV | 15% | 202 | 37% | 13 | 15% | 189 |  | <0.001 |
| **HAV vaccination history (≥1 dose)** |  |  |  |  |  |  |  |  |
| No | 32% | 430 | 23% | 8 | 33% | 422 |  |  |
| Yes | 68% | 903 | 77% | 27 | 67% | 876 |  | 0.228 |
| **HBV vaccination history (≥1 dose)** |  |  |  |  |  |  |  |  |
| No | 28% | 379 | 14% | 5 | 29% | 374 |  |  |
| Yes | 72% | 954 | 86% | 30 | 71% | 924 |  | 0.060 |
| **HPV vaccination history (≥1 dose)** |  |  |  |  |  |  |  |  |
| No | 58% | 771 | 26% | 9 | 59% | 762 |  |  |
| Yes | 42% | 562 | 74% | 26 | 41% | 536 |  | <0.001 |
| **SHS visit in last year** |  |  |  |  |  |  |  |  |
| No | 36% | 478 | 6% | 2 | 37% | 476 |  |  |
| Yes | 64% | 855 | 94% | 33 | 63% | 822 |  | <0.001 |
| **STI tested in last year** |  |  |  |  |  |  |  |  |
| No | 30% | 405 | 3% | 1 | 31% | 404 |  |  |
| Yes | 70% | 928 | 97% | 34 | 69% | 894 |  | <0.001 |
| **STI test positivity since August 2022** |  |  |  |  |  |  |  |  |
| Never STI tested | 9% | 123 | 3% | 1 | 9% | 122 |  |  |
| Tested before August 2022 | 38% | 513 | 20% | 7 | 39% | 506 |  |  |
| All negative/unknown recent test results since August 2022 | 41% | 541 | 34% | 12 | 41% | 529 |  |  |
| ≥1 recent positive test results since August 2022 | 12% | 156 | 43% | 15 | 11% | 141 |  | <0.001 |
| **Recreational drug use associated with chemsex in last year†** |  |  |  |  |  |  |  |  |
| No | 92% | 1,222 | 77% | 27 | 92% | 1,195 |  |  |
| Yes | 8% | 111 | 23% | 8 | 8% | 103 |  | 0.002 |
| **Number of recent male physical sex partners since August 2022** |  |  |  |  |  |  |  |  |
| None | 8% | 100 | 3% | 1 | 8% | 99 |  |  |
| 1 | 14% | 180 | 6% | 2 | 14% | 178 |  |  |
| 2 to 4 | 29% | 389 | 26% | 9 | 29% | 380 |  |  |
| 5 to 9 | 21% | 280 | 17% | 6 | 21% | 274 |  |  |
| ≥10 | 29% | 384 | 49% | 17 | 28% | 367 |  | 0.094 |
| **Met recent male physical sex partners in sex on premises venue, sex party, or PSE since August 2022** |  |  |  |  |  |  |  |  |
| No | 67% | 888 | 49% | 17 | 67% | 871 |  |  |
| Yes | 33% | 445 | 51% | 18 | 33% | 427 |  | 0.022 |
| **PrEP use since December 2021 (HIV negative/unknown status, n=1,131)¶** |  |  |  |  |  |  |  |  |
| No | 47% | 534 | 9% | 2 | 48% | 532 |  |  |
| Yes | 53% | 597 | 91% | 20 | 52% | 577 |  | <0.001 |
| **Behaviour modification since May 2022 to avoid getting mpox\*\*** |  |  |  |  |  |  |  |  |
| No | 47% | 626 | 51% | 18 | 47% | 608 |  |  |
| Yes | 53% | 707 | 49% | 17 | 53% | 690 |  | 0.592 |

\*Participants with self-reported mpox test positivity. Those without mpox test positivity (i.e. no diagnosis history) include those without a known positive test (n=67) and those without a test history (n=1231). †Includes those identifying as bisexual, straight, or another way. §Top two quartiles ("I am comfortable"/"I am very comfortable" from the question, "How would you best describe your current financial situation". ‡Includes crystal methamphetamine, mephedrone or gamma-hydroxybutyrate/gamma-butyrolactone. ¶Excludes people living with HIV (n=202). \*\*Behaviour modification includes self-report of any of the following from May 2022: fewer sexual partners, reduced visits to sex on premises venues or PSE (i.e. cruising grounds), and avoiding: all sex, condomless anal sex, skin-to-skin contact, or visiting clubs or crowds. HAV=Hepatitis A virus. HBV=Hepatitis B virus. HPV=Human papilloma virus. SHS=Sexual health service. STI=Sexually transmitted infection. PSE=Public sex environment (i.e. cruising grounds). PrEP=HIV pre-exposure prophylaxis. Percentages may not total 100 due to rounding.

**Table 2.**
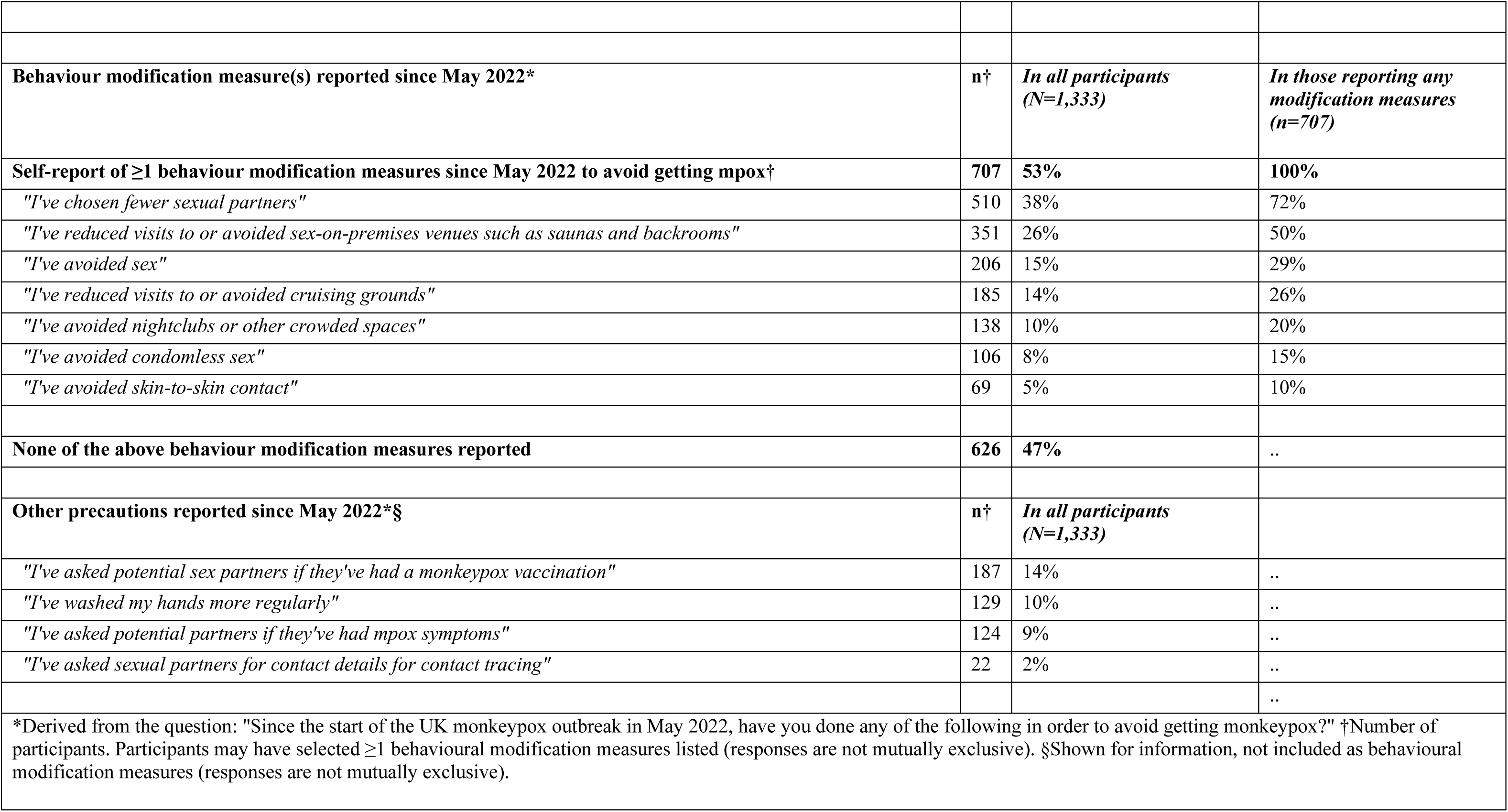
Behaviour modification measures and other precautions reported since May 2022 to avoid getting mpox by RiiSH-Mpox participants, November/December 2022.

### Mpox diagnosis history

Among all participants, 2.6% (95% CI: 1.8%-3.6%; 35/1,333) reported a mpox diagnosis history. In our sensitivity analysis, an additional 17 participants without mpox test positivity (n=3) or test history (n=14) reported self-perceived mpox, bringing a total of 52 participants with either mpox test positivity or self-perceived mpox (3.9% [95% CI: 2.9%-5.1%]) (Appendix II, Appendix IV).

Those with a mpox diagnosis history (n=35), compared to those without (n=1,298), were more likely to be older (91% vs 76% aged ≥35, p=0.031), born outside of the UK (37% vs 16%, p=0.001), report a comfortable financial situation (66% vs 48%, p=0.034), and living with HIV (37% vs 15%, p=<0.001). Higher levels of sexual health clinic engagement in the last year, recent STI test positivity, and sexual risk behaviours such as meeting partners at sex on premises venue, sex party, or PSE, were also reported in those with a mpox diagnosis history (Table 1). Both groups reported similar proportions of behaviour modification since May 2022 as a result of the mpox outbreak (49% vs 53%, p=0.592). These findings were similar to those found on the sensitivity analysis (Appendix IV).

### Mpox vaccination uptake

Over half of participants were offered a mpox vaccination by a SHS (58%, 771/1,333); vaccination uptake was 52% (95% CI: 49%-55%; 692/1,333) among all participants, or 90% (95% CI: 87%-92%; 692/771) among those offered a vaccine (Table 3, Appendix III). Less than half of those reporting vaccination had received a second dose (41%, 288/692).

**Table 3.** Mpox vaccine offer and uptake among RiiSH-Mpox participants by mpox vaccine eligibility, November/December 2022.

|  |  |  |  |  |  |  |
| --- | --- | --- | --- | --- | --- | --- |
|  |  | **Mpox vaccine offer and vaccination\*** |  |  | **Mpox vaccination uptake** |  |
|  | *Total offered vaccine* | *a) Offered and vaccinated* | *b) Offered but not vaccinated* | *c) Not offered and not vaccinated* | *In those offered vaccine†* | *In all participants‡* |
| Eligibility group | *n (row %)* | *n (row %)* | *n (row %)* | *n (row %)* | *% (95% CI)* | *% (95% CI)* |
| Eligible* for mpox vaccination (n=875) | 655 (75%) | 601 (69%) | 54 (6%) § | 220 (25%)** | 92% (89%-94%) | 69% (65%-72%) |
| Not eligible for mpox vaccination (n=458) | 116 (25%) | 91 (20%) | 25 (5%) | 342 (75%) | 78% (70%-86%) | 20% (16%-24%) |
| All RiiSH-Mpox participants (N=1,333) | 771 (75%) | 692 (52%) | 79 (6%) ¶ | 562 (42%)†† | 90% (88%-92%) | 52% (49%-55%) |
| *Self-reporting any of the following: meeting recent male physical sex partners at sex on premises venues, sex parties, or PSE (i.e. cruising grounds) (since August 2022) OR ≥10 recent male physical sex partners (since August 2022) OR recreational drug use associated with chemsex (in the last year) OR recent positive STI test (since August 2022) OR current PrEP use (since December 2021); †Percentage (%) reporting uptake of ≥1 mpox vaccine dose(s) among those offered an mpox vaccine $[(a/(a+b)) \times 100\%]$. ‡Percentage (%) reporting uptake of ≥1 mpox vaccine dose(s) among all group $[(a/(a+b+c)) \times 100\%]$. § Of participants mpox vaccine eligible and offered a vaccine, but not vaccinated (n=54): 28 (52%) were waiting to get vaccinated, 26 (48%) decided not to get vaccinated. ¶ Of all participants offered a mpox vaccine, but not vaccinated (n=79): 42 (53%) were waiting to get vaccinated, 37 (47%) decided not to get vaccinated. **Of those eligible and not offered, 38% (83/220) had visited an SHS since August 2022 and 57% (126/220) had visited an SHS in the last year. ††Of those not offered, 20% (111/562) had visited an SHS since August 2022 and 32% (182/562) had visited an SHS in the last year. 95% CI=95% confidence interval. SHS=Sexual health service. Percentages may not total 100 due to rounding. |  |  |  |  |  |  |

Of GBMSM considered vaccine eligible (66%, 875/1,333), three-quarters (75%, 655/875) were offered a vaccine (Table 3, Appendix III). Vaccination uptake was 69% (95% CI: 65%-72%; 601/875) among all eligible participants, or 92% (95% CI: 89%-94%; 601/655) in those eligible and offered a vaccine. In those not vaccinated but offered a vaccine (n=54), 48% (26/54) reported the decision not to get vaccinated (4.0% [26/655] of all eligible and offered), while a quarter of those eligible (25%, 220/875) were not offered a mpox vaccine. Second doses were reported by 41% (252/601) of eligible GBMSM vaccinated.

In sensitivity analyses using a lower threshold for multiple partners (>5) to assume vaccine eligibility, there was a similar level of uptake (Appendix VI).

### Factors associated with vaccination

Bivariate and multivariable associations with mpox vaccination uptake are presented in Table 4. We found evidence of bivariate association with mpox vaccination by age-group, sexual orientation, educational qualifications, employment, and current financial situation. In adjusted models, bisexual men (32% vs 54% in gay/homosexual men; aOR: 0.43, 95% CI: 0.29-0.62), those with education qualifications below degree-level (40% vs 59% in degree-level or higher; aOR: 0.50, 95% CI: 0.39-0.63), and those not employed (37% vs 55% in those employed; aOR: 0.59, 95% CI: 0.43-0.80) were less likely to report mpox vaccination. Participants reporting a single relationship status were more likely to be vaccinated (54% vs 50% of those in a relationship; aOR: 1.27, 95% CI: 1.01-1.60). We found no evidence of independent association to age, though lowest levels of vaccination were seen in those aged 16-24 years (30% vs 58% in those aged 45-54 years; aOR: 0.47, 95% CI: 0.25-0.87); those aged 16-24 years comprised only 5% of those mpox vaccinated. Reporting a positive STI test since August 2022 (aOR: 4.09, 95% CI: 2.69-6.22), having a HAV, HBV, or HPV vaccination history (aOR: 5.27, 95% CI: 3.72-7.47), reporting higher numbers of physical sex partners since August 2022 (aOR: 7.73, 95% CI: 5.06-11.8, ≥10 partners vs 1 partner), and reporting PrEP use since December 2021 (aOR: 7.09, 95% CI: 5.49-9.15) were the greatest individual predictors of mpox vaccination following sociodemographic adjustment. Among mpox vaccinated GBMSM, 87% (601/692) met proxy mpox vaccination eligibility. Those who met mpox vaccination eligibility were 8-times more likely to have been vaccinated relative to those who did not (aOR: 8.38, 95% CI: 6.35-11.1).

**Table 4.**
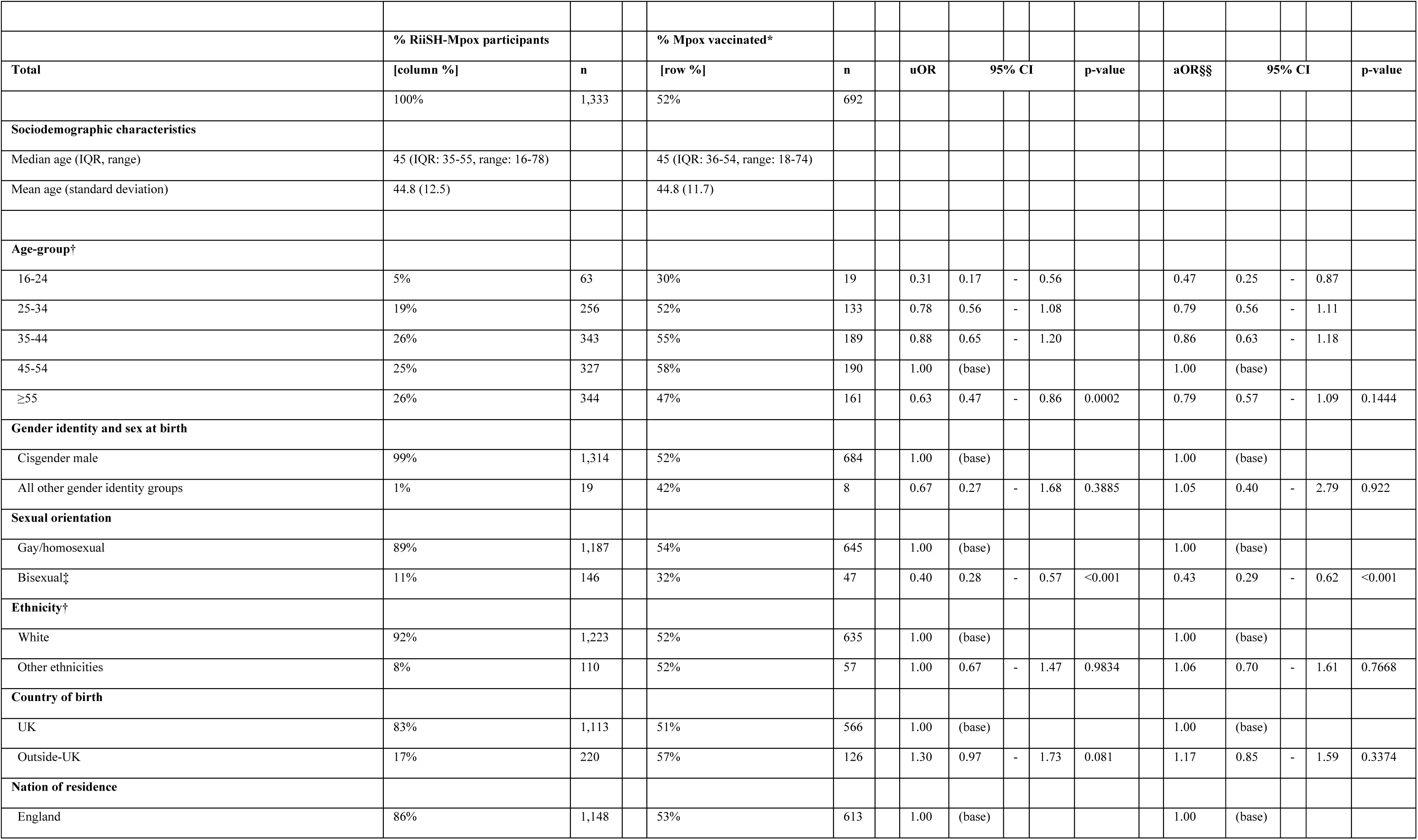
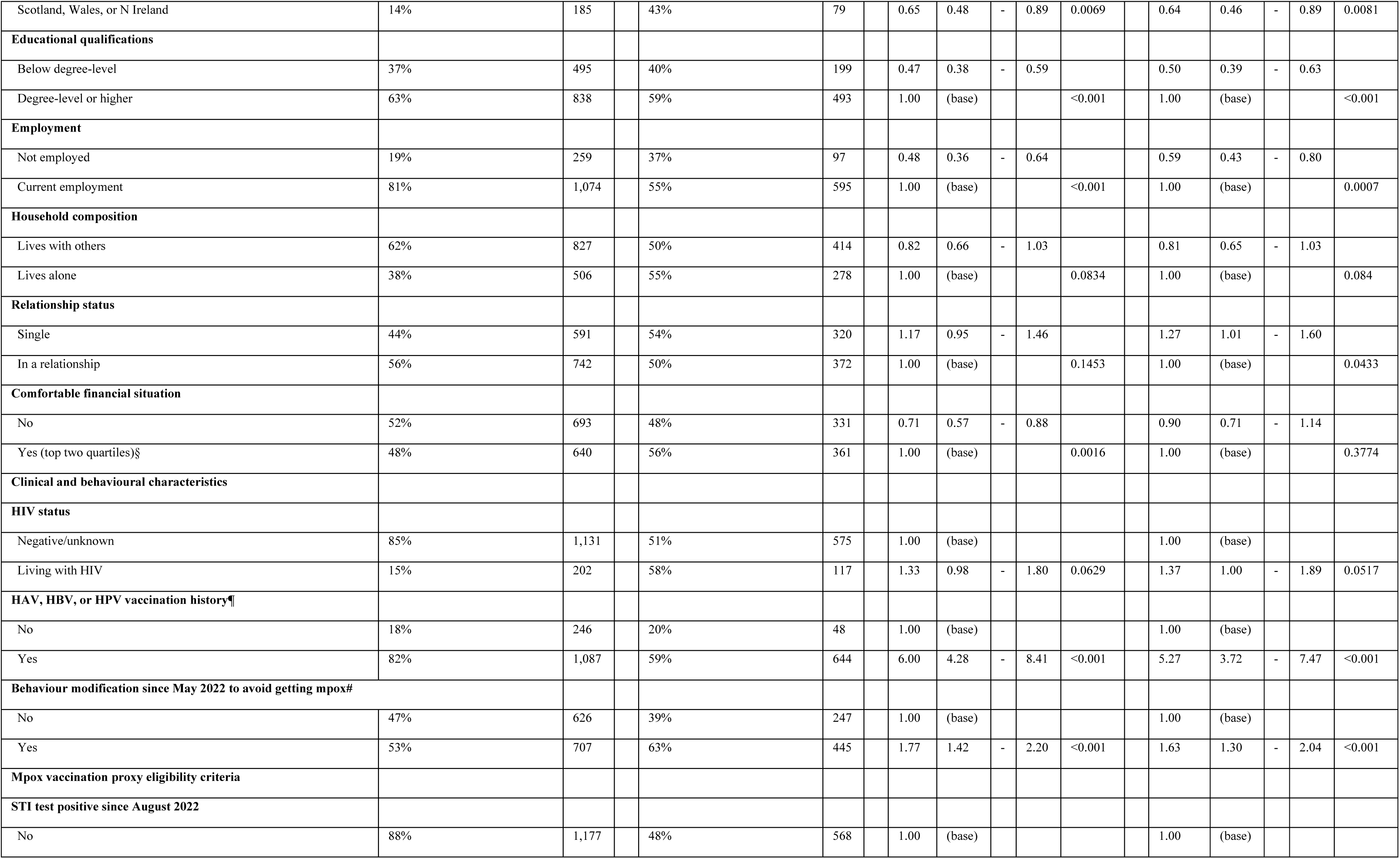
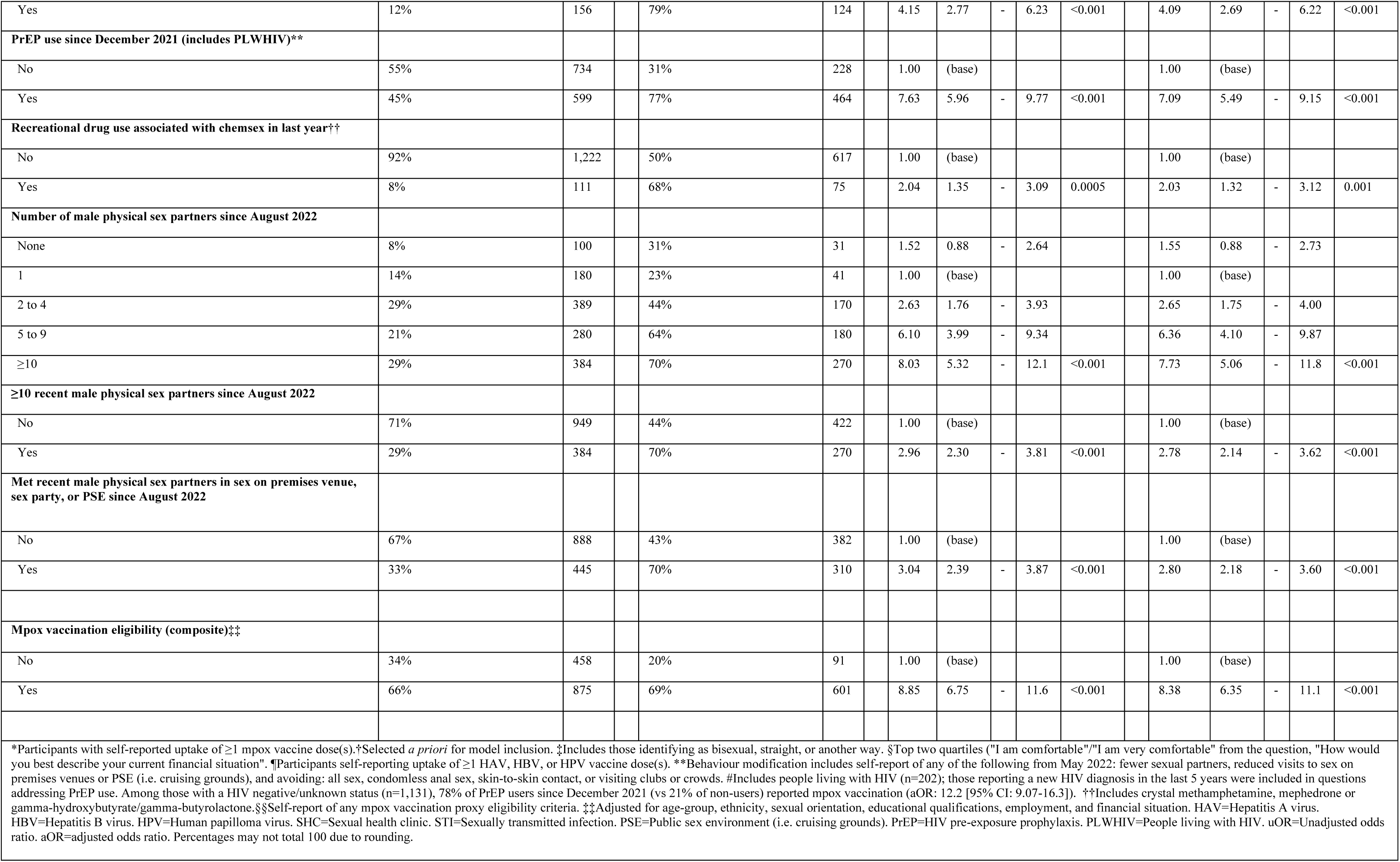
Characteristics of, and factors associated with self-reported mpox vaccination uptake among RiiSH-Mpox participants, November/December 2022.

|  |  |  |  |  |  |  |  |  |  |  |  |  |  |  |  |
| --- | --- | --- | --- | --- | --- | --- | --- | --- | --- | --- | --- | --- | --- | --- | --- |
|  | % RiiSH-Mpox participants |  | % Mpox vaccinated* |  |  |  |  |  |  |  |  |  |  |  |  |
| Total | [column %] | n | [row %] | n | uOR | 95% CI |  |  | p-value |  | aOR§§ | 95% CI |  |  | p-value |
|  | 100% | 1,333 | 52% | 692 |  |  |  |  |  |  |  |  |  |  |  |
| **Sociodemographic characteristics** |  |  |  |  |  |  |  |  |  |  |  |  |  |  |  |
| Median age (IQR, range) | 45 (IQR: 35-55, range: 16-78) |  | 45 (IQR: 36-54, range: 18-74) |  |  |  |  |  |  |  |  |  |  |  |  |
| Mean age (standard deviation) | 44.8 (12.5) |  | 44.8 (11.7) |  |  |  |  |  |  |  |  |  |  |  |  |
| **Age-group†** |  |  |  |  |  |  |  |  |  |  |  |  |  |  |  |
| 16-24 | 5% | 63 | 30% | 19 | 0.31 | 0.17 | - | 0.56 |  |  | 0.47 | 0.25 | - | 0.87 |  |
| 25-34 | 19% | 256 | 52% | 133 | 0.78 | 0.56 | - | 1.08 |  |  | 0.79 | 0.56 | - | 1.11 |  |
| 35-44 | 26% | 343 | 55% | 189 | 0.88 | 0.65 | - | 1.20 |  |  | 0.86 | 0.63 | - | 1.18 |  |
| 45-54 | 25% | 327 | 58% | 190 | 1.00 | (base) |  |  |  |  | 1.00 | (base) |  |  |  |
| ≥55 | 26% | 344 | 47% | 161 | 0.63 | 0.47 | - | 0.86 | 0.0002 |  | 0.79 | 0.57 | - | 1.09 | 0.1444 |
| **Gender identity and sex at birth** |  |  |  |  |  |  |  |  |  |  |  |  |  |  |  |
| Cisgender male | 99% | 1,314 | 52% | 684 | 1.00 | (base) |  |  |  |  | 1.00 | (base) |  |  |  |
| All other gender identity groups | 1% | 19 | 42% | 8 | 0.67 | 0.27 | - | 1.68 | 0.3885 |  | 1.05 | 0.40 | - | 2.79 | 0.922 |
| **Sexual orientation** |  |  |  |  |  |  |  |  |  |  |  |  |  |  |  |
| Gay/homosexual | 89% | 1,187 | 54% | 645 | 1.00 | (base) |  |  |  |  | 1.00 | (base) |  |  |  |
| Bisexual‡ | 11% | 146 | 32% | 47 | 0.40 | 0.28 | - | 0.57 | <0.001 |  | 0.43 | 0.29 | - | 0.62 | <0.001 |
| **Ethnicity†** |  |  |  |  |  |  |  |  |  |  |  |  |  |  |  |
| White | 92% | 1,223 | 52% | 635 | 1.00 | (base) |  |  |  |  | 1.00 | (base) |  |  |  |
| Other ethnicities | 8% | 110 | 52% | 57 | 1.00 | 0.67 | - | 1.47 | 0.9834 |  | 1.06 | 0.70 | - | 1.61 | 0.7668 |
| **Country of birth** |  |  |  |  |  |  |  |  |  |  |  |  |  |  |  |
| UK | 83% | 1,113 | 51% | 566 | 1.00 | (base) |  |  |  |  | 1.00 | (base) |  |  |  |
| Outside-UK | 17% | 220 | 57% | 126 | 1.30 | 0.97 | - | 1.73 | 0.081 |  | 1.17 | 0.85 | - | 1.59 | 0.3374 |
| **Nation of residence** |  |  |  |  |  |  |  |  |  |  |  |  |  |  |  |
| England | 86% | 1,148 | 53% | 613 | 1.00 | (base) |  |  |  |  | 1.00 | (base) |  |  |  |

|  |  |  |  |  |  |  |  |  |  |  |  |  |  |  |  |  |  |
| --- | --- | --- | --- | --- | --- | --- | --- | --- | --- | --- | --- | --- | --- | --- | --- | --- | --- |
| Scotland, Wales, or N Ireland | 14% | 185 |  | 43% | 79 |  | 0.65 | 0.48 | - | 0.89 | 0.0069 |  | 0.64 | 0.46 | - | 0.89 | 0.0081 |
| **Educational qualifications** |  |  |  |  |  |  |  |  |  |  |  |  |  |  |  |  |  |
| Below degree-level | 37% | 495 |  | 40% | 199 |  | 0.47 | 0.38 | - | 0.59 |  |  | 0.50 | 0.39 | - | 0.63 |  |
| Degree-level or higher | 63% | 838 |  | 59% | 493 |  | 1.00 | (base) |  |  | <0.001 |  | 1.00 | (base) |  |  | <0.001 |
| **Employment** |  |  |  |  |  |  |  |  |  |  |  |  |  |  |  |  |  |
| Not employed | 19% | 259 |  | 37% | 97 |  | 0.48 | 0.36 | - | 0.64 |  |  | 0.59 | 0.43 | - | 0.80 |  |
| Current employment | 81% | 1,074 |  | 55% | 595 |  | 1.00 | (base) |  |  | <0.001 |  | 1.00 | (base) |  |  | 0.0007 |
| **Household composition** |  |  |  |  |  |  |  |  |  |  |  |  |  |  |  |  |  |
| Lives with others | 62% | 827 |  | 50% | 414 |  | 0.82 | 0.66 | - | 1.03 |  |  | 0.81 | 0.65 | - | 1.03 |  |
| Lives alone | 38% | 506 |  | 55% | 278 |  | 1.00 | (base) |  |  | 0.0834 |  | 1.00 | (base) |  |  | 0.084 |
| **Relationship status** |  |  |  |  |  |  |  |  |  |  |  |  |  |  |  |  |  |
| Single | 44% | 591 |  | 54% | 320 |  | 1.17 | 0.95 | - | 1.46 |  |  | 1.27 | 1.01 | - | 1.60 |  |
| In a relationship | 56% | 742 |  | 50% | 372 |  | 1.00 | (base) |  |  | 0.1453 |  | 1.00 | (base) |  |  | 0.0433 |
| **Comfortable financial situation** |  |  |  |  |  |  |  |  |  |  |  |  |  |  |  |  |  |
| No | 52% | 693 |  | 48% | 331 |  | 0.71 | 0.57 | - | 0.88 |  |  | 0.90 | 0.71 | - | 1.14 |  |
| Yes (top two quartiles)§ | 48% | 640 |  | 56% | 361 |  | 1.00 | (base) |  |  | 0.0016 |  | 1.00 | (base) |  |  | 0.3774 |
| **Clinical and behavioural characteristics** |  |  |  |  |  |  |  |  |  |  |  |  |  |  |  |  |  |
| **HIV status** |  |  |  |  |  |  |  |  |  |  |  |  |  |  |  |  |  |
| Negative/unknown | 85% | 1,131 |  | 51% | 575 |  | 1.00 | (base) |  |  |  |  | 1.00 | (base) |  |  |  |
| Living with HIV | 15% | 202 |  | 58% | 117 |  | 1.33 | 0.98 | - | 1.80 | 0.0629 |  | 1.37 | 1.00 | - | 1.89 | 0.0517 |
| **HAV, HBV, or HPV vaccination history¶** |  |  |  |  |  |  |  |  |  |  |  |  |  |  |  |  |  |
| No | 18% | 246 |  | 20% | 48 |  | 1.00 | (base) |  |  |  |  | 1.00 | (base) |  |  |  |
| Yes | 82% | 1,087 |  | 59% | 644 |  | 6.00 | 4.28 | - | 8.41 | <0.001 |  | 5.27 | 3.72 | - | 7.47 | <0.001 |
| **Behaviour modification since May 2022 to avoid getting mpox#** |  |  |  |  |  |  |  |  |  |  |  |  |  |  |  |  |  |
| No | 47% | 626 |  | 39% | 247 |  | 1.00 | (base) |  |  |  |  | 1.00 | (base) |  |  |  |
| Yes | 53% | 707 |  | 63% | 445 |  | 1.77 | 1.42 | - | 2.20 | <0.001 |  | 1.63 | 1.30 | - | 2.04 | <0.001 |
| **Mpox vaccination proxy eligibility criteria** |  |  |  |  |  |  |  |  |  |  |  |  |  |  |  |  |  |
| **STI test positive since August 2022** |  |  |  |  |  |  |  |  |  |  |  |  |  |  |  |  |  |
| No | 88% | 1,177 |  | 48% | 568 |  | 1.00 | (base) |  |  |  |  | 1.00 | (base) |  |  |  |

|  |  |  |  |  |  |  |  |  |  |  |  |  |  |  |
| --- | --- | --- | --- | --- | --- | --- | --- | --- | --- | --- | --- | --- | --- | --- |
| Yes | 12% | 156 | 79% | 124 | 4.15 | 2.77 | - | 6.23 | <0.001 | 4.09 | 2.69 | - | 6.22 | <0.001 |
| **PrEP use since December 2021 (includes PLWHIV)\*\*** |  |  |  |  |  |  |  |  |  |  |  |  |  |  |
| No | 55% | 734 | 31% | 228 | 1.00 | (base) |  |  |  | 1.00 | (base) |  |  |  |
| Yes | 45% | 599 | 77% | 464 | 7.63 | 5.96 | - | 9.77 | <0.001 | 7.09 | 5.49 | - | 9.15 | <0.001 |
| **Recreational drug use associated with chemsex in last year††** |  |  |  |  |  |  |  |  |  |  |  |  |  |  |
| No | 92% | 1,222 | 50% | 617 | 1.00 | (base) |  |  |  | 1.00 | (base) |  |  |  |
| Yes | 8% | 111 | 68% | 75 | 2.04 | 1.35 | - | 3.09 | 0.0005 | 2.03 | 1.32 | - | 3.12 | 0.001 |
| **Number of male physical sex partners since August 2022** |  |  |  |  |  |  |  |  |  |  |  |  |  |  |
| None | 8% | 100 | 31% | 31 | 1.52 | 0.88 | - | 2.64 |  | 1.55 | 0.88 | - | 2.73 |  |
| 1 | 14% | 180 | 23% | 41 | 1.00 | (base) |  |  |  | 1.00 | (base) |  |  |  |
| 2 to 4 | 29% | 389 | 44% | 170 | 2.63 | 1.76 | - | 3.93 |  | 2.65 | 1.75 | - | 4.00 |  |
| 5 to 9 | 21% | 280 | 64% | 180 | 6.10 | 3.99 | - | 9.34 |  | 6.36 | 4.10 | - | 9.87 |  |
| ≥10 | 29% | 384 | 70% | 270 | 8.03 | 5.32 | - | 12.1 | <0.001 | 7.73 | 5.06 | - | 11.8 | <0.001 |
| **≥10 recent male physical sex partners since August 2022** |  |  |  |  |  |  |  |  |  |  |  |  |  |  |
| No | 71% | 949 | 44% | 422 | 1.00 | (base) |  |  |  | 1.00 | (base) |  |  |  |
| Yes | 29% | 384 | 70% | 270 | 2.96 | 2.30 | - | 3.81 | <0.001 | 2.78 | 2.14 | - | 3.62 | <0.001 |
| **Met recent male physical sex partners in sex on premises venue, sex party, or PSE since August 2022** |  |  |  |  |  |  |  |  |  |  |  |  |  |  |
| No | 67% | 888 | 43% | 382 | 1.00 | (base) |  |  |  | 1.00 | (base) |  |  |  |
| Yes | 33% | 445 | 70% | 310 | 3.04 | 2.39 | - | 3.87 | <0.001 | 2.80 | 2.18 | - | 3.60 | <0.001 |
| **Mpox vaccination eligibility (composite)‡‡** |  |  |  |  |  |  |  |  |  |  |  |  |  |  |
| No | 34% | 458 | 20% | 91 | 1.00 | (base) |  |  |  | 1.00 | (base) |  |  |  |
| Yes | 66% | 875 | 69% | 601 | 8.85 | 6.75 | - | 11.6 | <0.001 | 8.38 | 6.35 | - | 11.1 | <0.001 |

In sensitivity analyses examining sociodemographic factors associated with mpox vaccination among those vaccine eligible, we found consistent findings to analyses among all participants, where mpox vaccine uptake was less likely in bisexual men (aOR: 0.49, 95% CI: 0.30-0.79 vs gay/homosexual men), those with lower educational qualifications (aOR: 0.46; 95% CI: 0.34-0.63), and those who were employed (aOR: 0.63; 95% CI: 0.42-0.95) (Appendix V).

## Discussion

In this large, online community survey of GBMSM in the UK undertaken shortly after the 2022 mpox outbreak, one in two of all participants reported adopting a risk modification measure and seven in 10 eligible participants had been mpox vaccinated, with very high uptake in those that were offered a vaccine. Most participants who were vaccinated met proxy eligibility criteria, and among all participants, vaccine uptake was associated with higher levels of sexual risk, suggesting fidelity to targeted vaccination set out by national guidelines during rapid vaccine roll-out across the UK in June 2022. Demographic and behavioural characteristics of those mpox test positive broadly reflected those described in enhanced surveillance of confirmed cases undertaken by UKHSA (10). Over a third of those who tested positive for mpox were living with HIV, consistent with mpox case reporting (20, 21).

To our knowledge, this is the first study examining mpox vaccination and self-reported behavioural modification among a community sample of GBMSM in the UK. We found mpox vaccine uptake was similar to levels reported among all and eligible (51%, 66%, respectively) transgender people and GBMSM sexual health clinic attendees shortly following vaccination implementation in British Columbia (22). Changes to sexual behaviour have been reported among GBMSM in the UK during periods of COVID-19 restrictions, however, there has been limited evidence exploring behavioural modification in response to the 2022 mpox outbreak. While sexual mixing outside of households generally persisted through COVID-19 pandemic waves (19, 23), studies noted swift rebounds in sexual risk behaviour following easing of social restrictions (18). Our findings support other evidence of behavioural modification among GBMSM during the mpox outbreak, as UK surveillance data show a concurrent, but temporary decline in lymphogranuloma venereum (LGV) and *Shigella* species, infections that primarily circulate in the dense, interconnected sexual networks associated with mpox transmission (10, 24).

In our study, lower vaccine uptake was reported in those with lower educational qualifications and those without employment; these differences were also found in sensitivity analyses restricted to those considered vaccine eligible. These findings mirror COVID-19 vaccine uptake inequalities found in the previous RiiSH survey where, during the time of that survey (December 2021), UK COVID-19 vaccination was widely accessible (25). We found bisexual and straight-identifying participants also had a lower likelihood of reporting mpox vaccination, consistent with findings in a smaller cross-sectional study exploring mpox vaccination uptake (22). The impact and epidemiology of mpox and associated morbidity in bisexual and heterosexual-identifying men who have sex with men (MSM) is unknown; a previous systematic review (26) reports lower sexual health service engagement despite high sexual risk in heterosexual-identifying MSM, suggesting a need for tailored vaccination promotion efforts for these groups in cases of mpox resurgence or endemicity, or with other sexually transmissible pathogens.

National guidance for targeting mpox vaccination recommended using markers of historical sexual risk shown to be associated with sexually transmitted infection (STI) and HIV incidence (27, 28). Most mpox vaccinated GBMSM met proxy eligibility criteria, however a third of those considered vaccine eligible did not report mpox vaccination, most of whom did not receive a vaccine offer. Findings could reflect SHS access barriers as only a third of those vaccine eligible and not offered a vaccine had visited a SHS since August 2022; in contrast, this may add to indications that some GBMSM do not regularly engage with SHS (29, 30). Vaccine provision and service-level constraints as a result of increased infection control measures needed for mpox management, as well as service reconfigurations, that included online triage, could have limited face-to-face offer and subsequent uptake (31, 32). To ensure anonymity of the respondents, the survey did not collect any data to indicate the region of residence of participants, where offer and uptake of vaccination may have been lower in regions of England that did not experience very large outbreaks or where local services did not provide vaccination. Individual-level barriers to vaccination, such as low perceived risk, may also have limited vaccine uptake among those eligible, especially once mpox incidence fell sharply across the UK after cases peaked in July.

Less than half of mpox-vaccinated participants reported already having received a second dose, despite only recent availability (September 2022) relative to survey completion, and is similar to the proportion reported nationally through May 2023 (11). Further work exploring low vaccine course completion is needed, especially as little data exist on the length of protection conferred following a single mpox vaccine dose and complete vaccination (33). Though rapid, first-dose vaccination provision has been recommended in the context of outbreak control following efficacy studies (16), since September 2022, UKHSA and the Joint Committee on Vaccination and Immunisation (JCVI) have recommended two dose offers in eligible groups (34), with the targeted mpox vaccination programme set to end in July 2023 (14).

### Limitations

The RiiSH-Mpox survey is part of a series of repeat, cross-sectional surveys using consistent methodology, providing key behavioural insights that supplement routine national surveillance data for STIs and HIV. This study represents one of the few, timely examinations of mpox in a community sample of GBMSM, contextualising interpretation of the trends in the mpox outbreak in the UK. However, our study is subject to key limitations due to its cross-sectional design. We cannot determine the time of vaccination in relation to most reported sexual risk behaviours comprising eligibility. Moreover, given the report of behavioural risk modification resulting from the mpox outbreak, vaccine eligibility could be underestimated, though behaviour could have changed following vaccine uptake. While observational studies of GBMSM in the UK and other high-income countries have reported a high willingness to be vaccinated against mpox (35–37), uptake estimates may be subject to sampling and social desirability bias. RiiSH-Mpox participants may represent a more health-literate sample, given higher educational attainment and employment relative to national probability survey estimates in GBMSM (38). Prior RiiSH cross-sectional samples have reported near universal uptake of complete COVID-19 vaccination (25). Though RiiSH-Mpox participants may not be representative of the wider GBMSM population in the UK, our study sample likely represents key groups targeted for mpox vaccination, in addition to vaccination for other sexually transmissible pathogens. Finally, small subgroup sizes across ethnicity and gender identity indicators limit assessment of inequalities in vaccine uptake.

## Conclusion

While high mpox vaccine uptake in eligible GBMSM and adoption of risk modification measures likely led to the reduction of mpox incidence in July 2022, the degree of contribution of each in the UK is unknown. Prior to, and during the reactive vaccine programme, timely vaccine resource information distributed and amplified by community-based organisations for GBMSM and other groups at risk likely contributed to the sharp drop in mpox incidence by the end of 2022 (8, 39).

In the UK, the use of key behavioural proxies guided mpox vaccination eligibility for GBMSM and facilitated vaccination implementation for those at risk of mpox. Any future targeted vaccination roll-out will need to consider rapid, yet equitable provision strategies, and must engage with the underserved via local outreach and community groups to maximise reach and vaccine uptake, and to minimise mpox-related stigma (40). Uptake barriers in sexual minorities, and those with lower social and financial capital, groups already described to have unmet sexual health need (18), must be understood to minimise exacerbation of vaccine uptake inequalities. Moreover, effective vaccination offer and provision strategies for those utilising online sexual health services warrant exploration as sexual health reconfigurations, often led by online service expansion, continue. Optimising SHS and outreach-led mpox vaccine offer, as part of a package of preventative interventions for those who may have unmet need, should be considered to maximise the benefit of each sexual health contact.

To reduce the likelihood of any future mpox outbreaks given threats of resurgence, provision of first mpox vaccination doses and completion of the course of vaccination in those who have received their first dose must be urgently prioritised.

## Contributions

QE, DP, RW, DJ, DR, GH, CHM, JS, HM, KS, HF, HCh, HCo, NOB, and KF reviewed and updated the survey instrument. Survey implementation, data collection and data management were carried out by QE, RW, and DR. DO, HM, and QE conceived analysis design with review and contributions from DP, RW, DJ, DR, GH, CHM, JS, KS, KF. DO conducted analyses and, with JRGB, wrote the first manuscript draft. All authors contributed to successive drafts and reviewed and approved the final manuscript.

## Ethics statement

Ethical approval of this study was provided by the UKHSA Research and Ethics Governance Group (REGG; ref: R&D 524). Online informed consent was received from all participants and all methods were performed in accordance with guidelines and regulations set by the UKHSA REGG.

## Competing interests

Authors have no competing interests to declare.

## Data availability statement

The data that support the findings of this study are available upon reasonable request from the UK Health Security Agency (UKHSA). Requests can be directed to Dr Hamish Mohammed.

## Funding

This study did not receive any funding and was conducted as part of the UKHSA public health response to mpox.

## Acknowledgements

The authors wish to thank all the participants who took part in this study. We acknowledge members of the National Institute for Health and Care Research Health Protection Research Unit (NIHR HPRU) in Blood Borne and Sexually Transmitted Infections (BBSTI) Steering Committee: Professor Caroline Sabin (HPRU Director), Dr John Saunders (UK Health Security Agency Lead), Professor Catherine Mercer, Dr Hamish Mohammed (previously Professor Gwenda Hughes), Professor Greta Rait, Dr Ruth Simmons, Professor William Rosenberg, Dr Tamyo Mbisa, Professor Rosalind Raine, Dr Sema Mandal, Dr Rosamund Yu, Dr Samreen Ijaz, Dr Fabiana Lorencatto, Dr Rachel Hunter, Dr Kirsty Foster and Dr Mamooma Tahir. The authors would also like to thank Takudzwa Mukiwa (Terrence Higgins Trust) and Will Nutland (The Love Tank) for their review of the mpox module in the RiiSH-Mpox survey used for this research.

---

**Appendix I:**
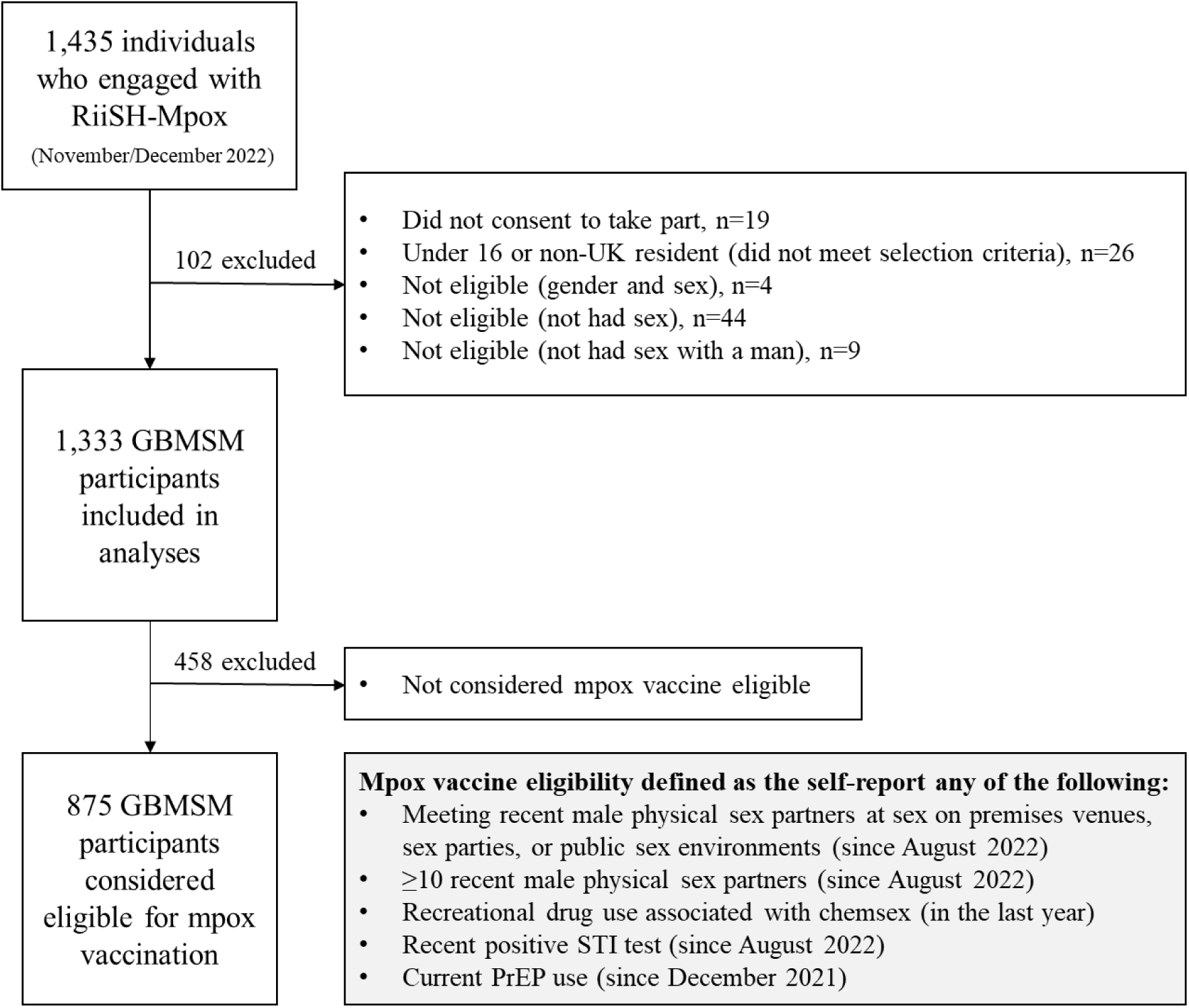
RiiSH-Mpox participant flowchart.

**Appendix II:**
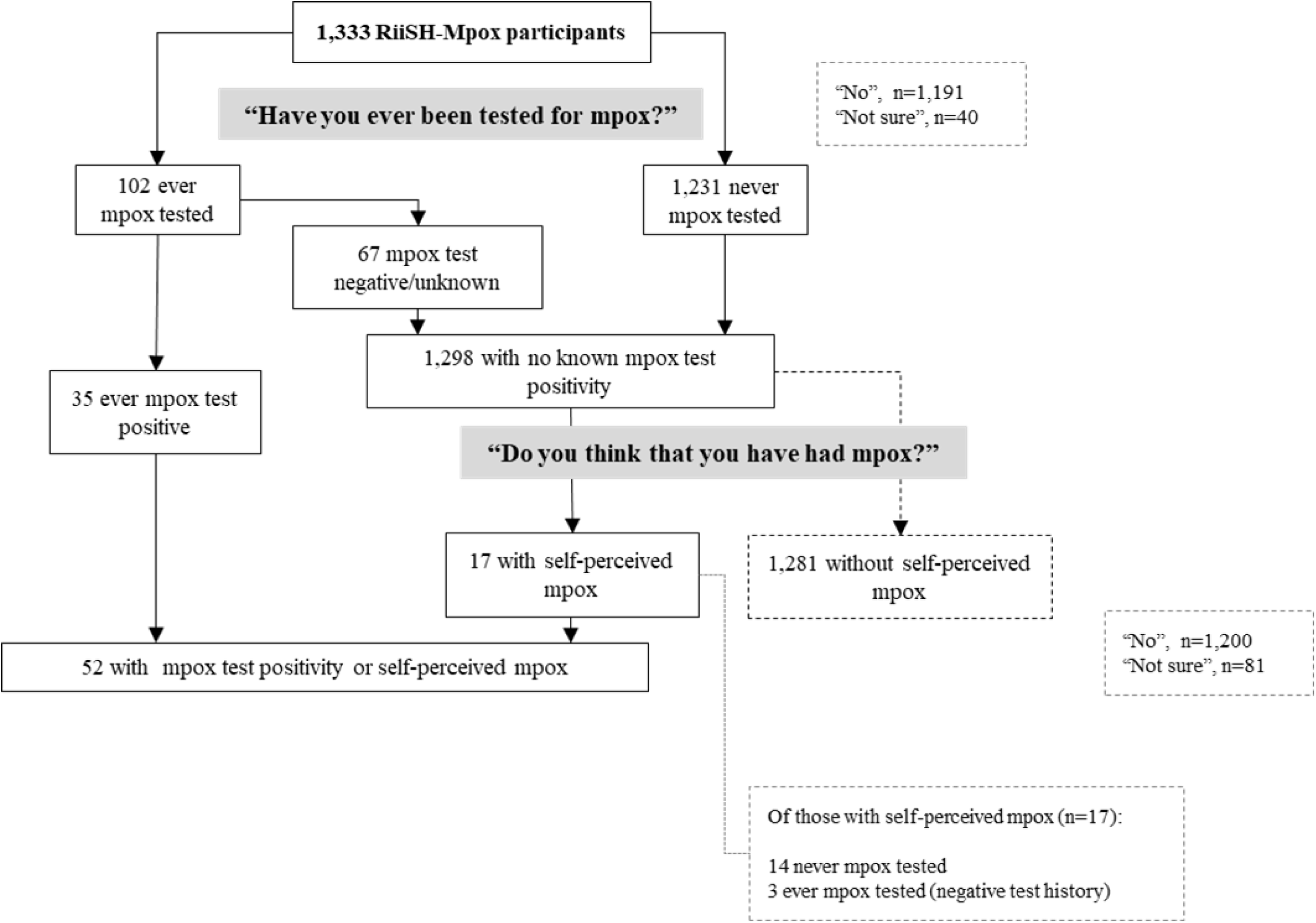
Self-reported mpox positivity and self-perceived mpox in RiiSH-Mpox participants, November/December 2022.

**Appendix III.**
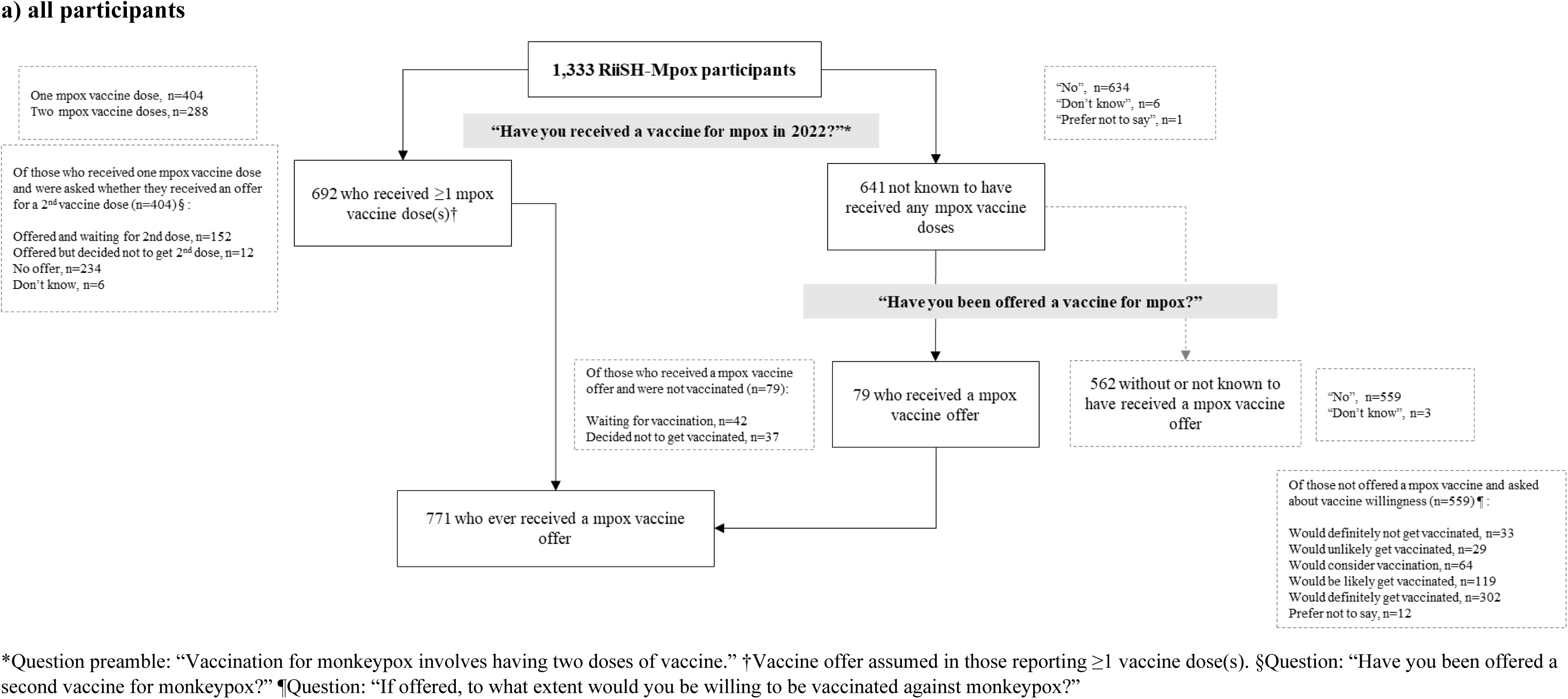
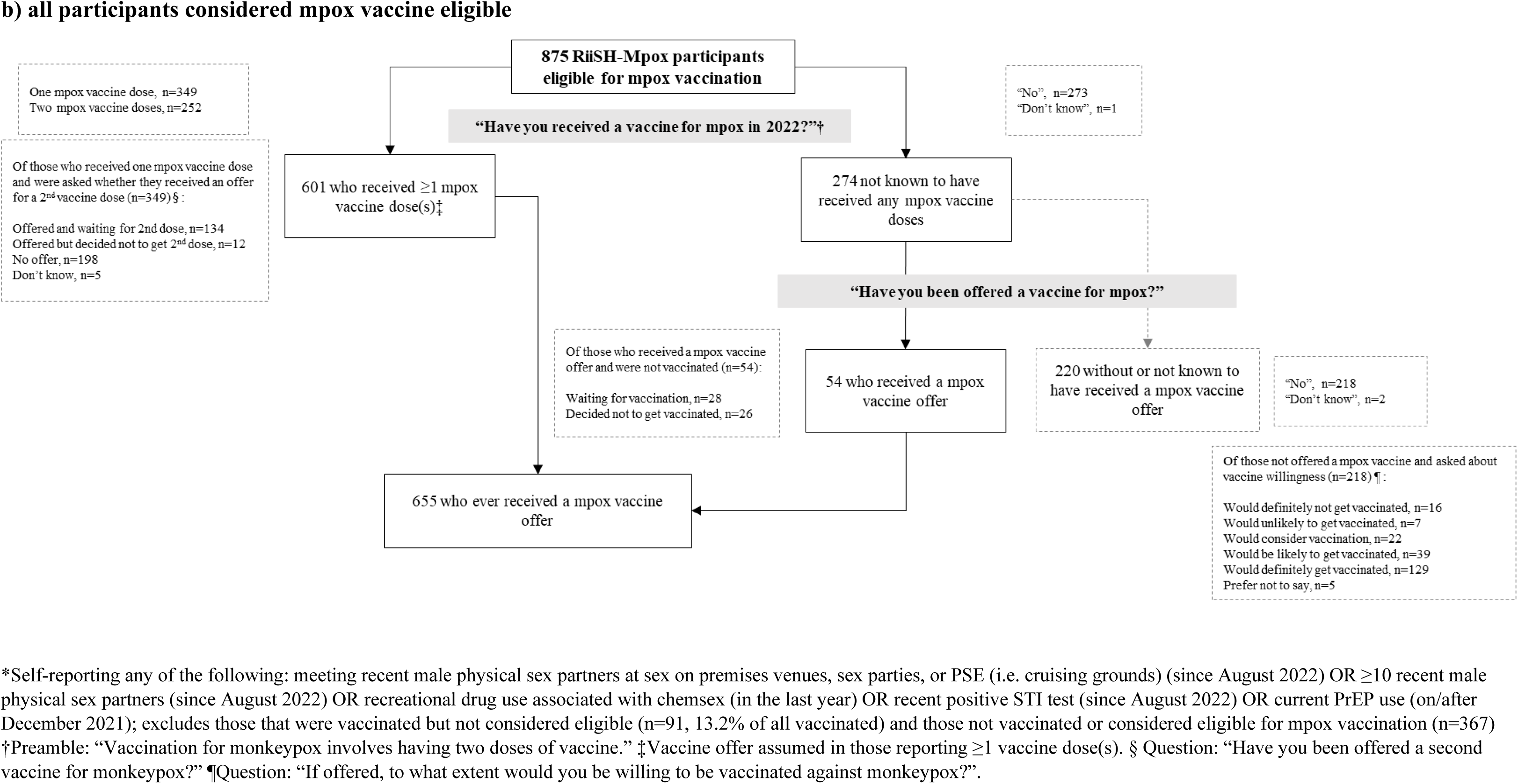
Mpox vaccination and offered reported in all RiiSH-Mpox participants and those considered mpox vaccine eligible, November/December 2022.

**Appendix IV.**
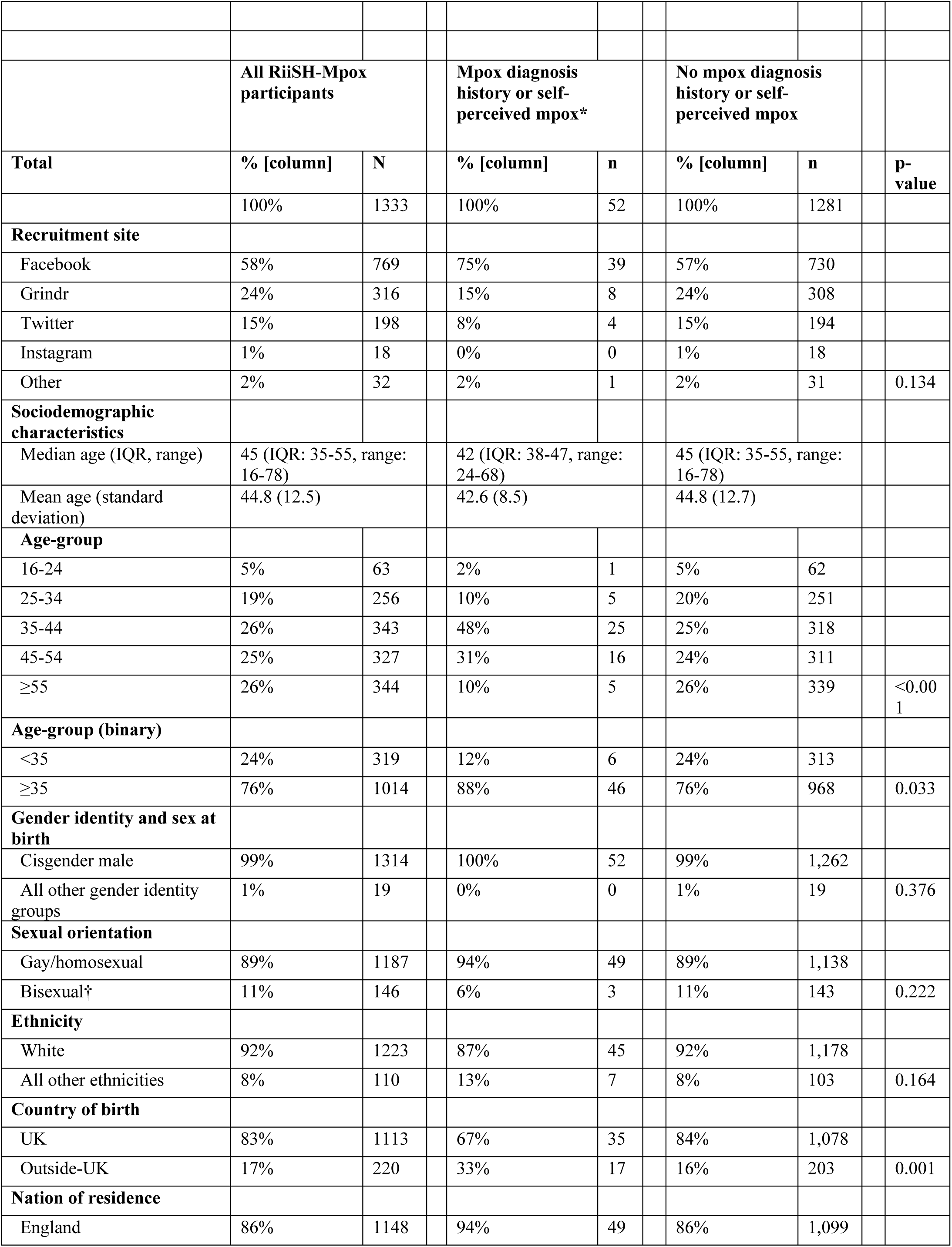
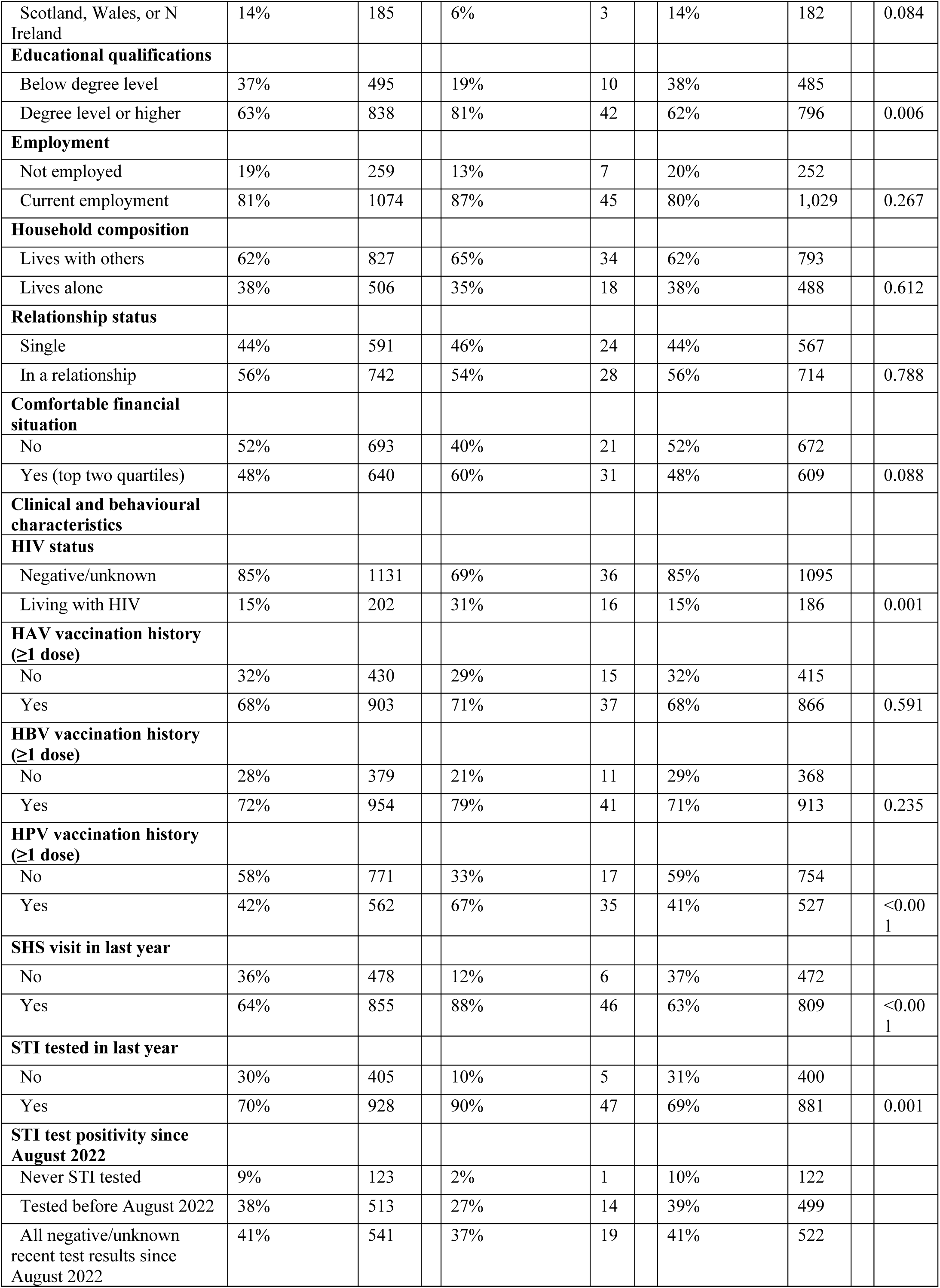
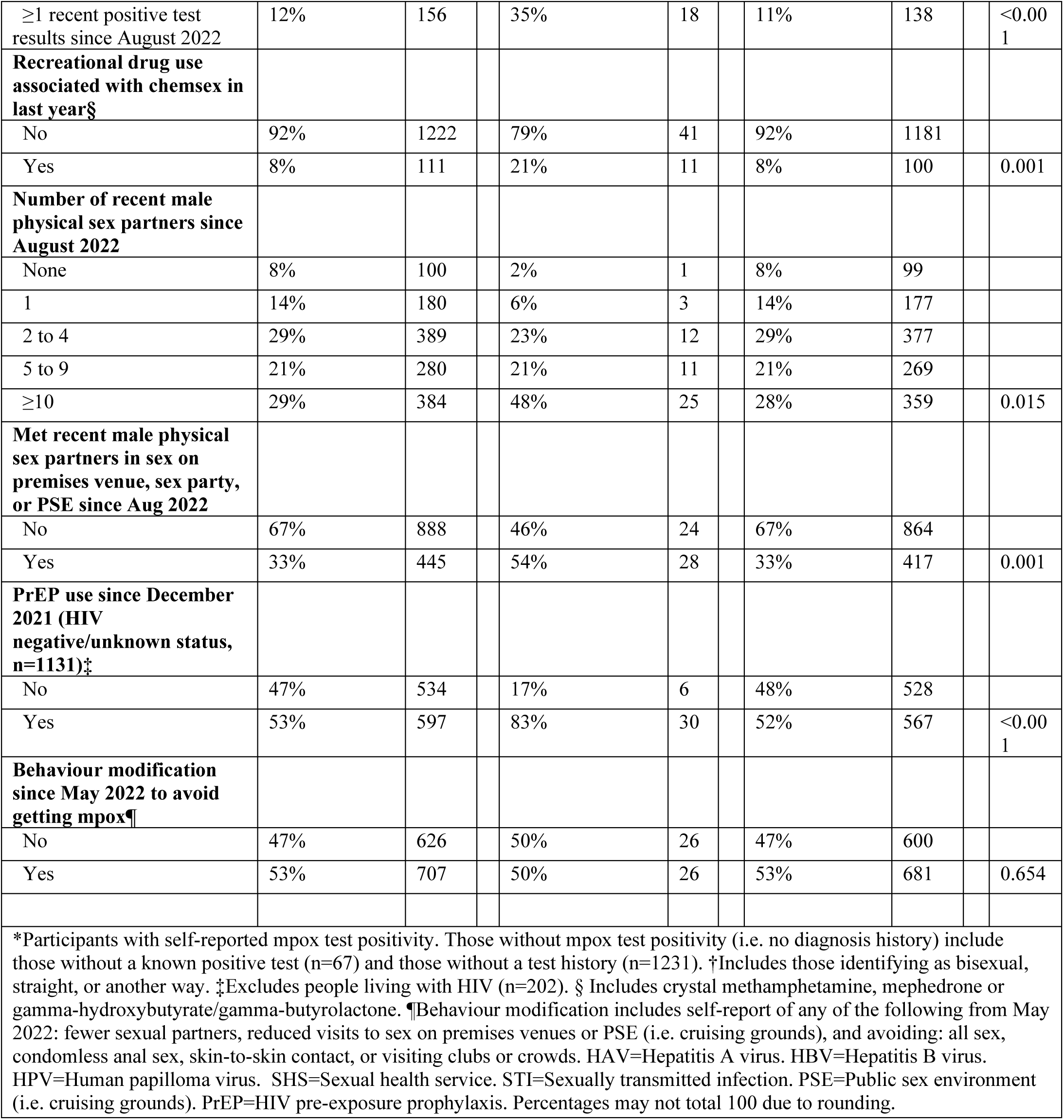
Sociodemographic and clinical characteristics of RiiSH-Mpox participants with and without a mpox diagnosis history that includes self-perceived mpox, November/December 2022.

|  |  |  |  |  |  |  |  |
| --- | --- | --- | --- | --- | --- | --- | --- |
|  | **All RiiSH-Mpox participants** |  | **Mpox diagnosis history or self-perceived mpox\*** |  | **No mpox diagnosis history or self-perceived mpox** |  |  |
| **Total** | **% [column]** | **N** | **% [column]** | **n** | **% [column]** | **n** | **p-value** |
|  | 100% | 1333 | 100% | 52 | 100% | 1281 |  |
| **Recruitment site** |  |  |  |  |  |  |  |
| Facebook | 58% | 769 | 75% | 39 | 57% | 730 |  |
| Grindr | 24% | 316 | 15% | 8 | 24% | 308 |  |
| Twitter | 15% | 198 | 8% | 4 | 15% | 194 |  |
| Instagram | 1% | 18 | 0% | 0 | 1% | 18 |  |
| Other | 2% | 32 | 2% | 1 | 2% | 31 | 0.134 |
| **Sociodemographic characteristics** |  |  |  |  |  |  |  |
| Median age (IQR, range) | 45 (IQR: 35-55, range: 16-78) |  | 42 (IQR: 38-47, range: 24-68) |  | 45 (IQR: 35-55, range: 16-78) |  |  |
| Mean age (standard deviation) | 44.8 (12.5) |  | 42.6 (8.5) |  | 44.8 (12.7) |  |  |
| **Age-group** |  |  |  |  |  |  |  |
| 16-24 | 5% | 63 | 2% | 1 | 5% | 62 |  |
| 25-34 | 19% | 256 | 10% | 5 | 20% | 251 |  |
| 35-44 | 26% | 343 | 48% | 25 | 25% | 318 |  |
| 45-54 | 25% | 327 | 31% | 16 | 24% | 311 |  |
| ≥55 | 26% | 344 | 10% | 5 | 26% | 339 | <0.001 |
| **Age-group (binary)** |  |  |  |  |  |  |  |
| <35 | 24% | 319 | 12% | 6 | 24% | 313 |  |
| ≥35 | 76% | 1014 | 88% | 46 | 76% | 968 | 0.033 |
| **Gender identity and sex at birth** |  |  |  |  |  |  |  |
| Cisgender male | 99% | 1314 | 100% | 52 | 99% | 1,262 |  |
| All other gender identity groups | 1% | 19 | 0% | 0 | 1% | 19 | 0.376 |
| **Sexual orientation** |  |  |  |  |  |  |  |
| Gay/homosexual | 89% | 1187 | 94% | 49 | 89% | 1,138 |  |
| Bisexual† | 11% | 146 | 6% | 3 | 11% | 143 | 0.222 |
| **Ethnicity** |  |  |  |  |  |  |  |
| White | 92% | 1223 | 87% | 45 | 92% | 1,178 |  |
| All other ethnicities | 8% | 110 | 13% | 7 | 8% | 103 | 0.164 |
| **Country of birth** |  |  |  |  |  |  |  |
| UK | 83% | 1113 | 67% | 35 | 84% | 1,078 |  |
| Outside-UK | 17% | 220 | 33% | 17 | 16% | 203 | 0.001 |
| **Nation of residence** |  |  |  |  |  |  |  |
| England | 86% | 1148 | 94% | 49 | 86% | 1,099 |  |
| Scotland, Wales, or N Ireland | 14% | 185 | 6% | 3 | 14% | 182 | 0.084 |
| **Educational qualifications** |  |  |  |  |  |  |  |
| Below degree level | 37% | 495 | 19% | 10 | 38% | 485 |  |
| Degree level or higher | 63% | 838 | 81% | 42 | 62% | 796 | 0.006 |
| **Employment** |  |  |  |  |  |  |  |
| Not employed | 19% | 259 | 13% | 7 | 20% | 252 |  |
| Current employment | 81% | 1074 | 87% | 45 | 80% | 1,029 | 0.267 |
| **Household composition** |  |  |  |  |  |  |  |
| Lives with others | 62% | 827 | 65% | 34 | 62% | 793 |  |
| Lives alone | 38% | 506 | 35% | 18 | 38% | 488 | 0.612 |
| **Relationship status** |  |  |  |  |  |  |  |
| Single | 44% | 591 | 46% | 24 | 44% | 567 |  |
| In a relationship | 56% | 742 | 54% | 28 | 56% | 714 | 0.788 |
| **Comfortable financial situation** |  |  |  |  |  |  |  |
| No | 52% | 693 | 40% | 21 | 52% | 672 |  |
| Yes (top two quartiles) | 48% | 640 | 60% | 31 | 48% | 609 | 0.088 |
| **Clinical and behavioural characteristics** |  |  |  |  |  |  |  |
| **HIV status** |  |  |  |  |  |  |  |
| Negative/unknown | 85% | 1131 | 69% | 36 | 85% | 1095 |  |
| Living with HIV | 15% | 202 | 31% | 16 | 15% | 186 | 0.001 |
| **HAV vaccination history (≥1 dose)** |  |  |  |  |  |  |  |
| No | 32% | 430 | 29% | 15 | 32% | 415 |  |
| Yes | 68% | 903 | 71% | 37 | 68% | 866 | 0.591 |
| **HBV vaccination history (≥1 dose)** |  |  |  |  |  |  |  |
| No | 28% | 379 | 21% | 11 | 29% | 368 |  |
| Yes | 72% | 954 | 79% | 41 | 71% | 913 | 0.235 |
| **HPV vaccination history (≥1 dose)** |  |  |  |  |  |  |  |
| No | 58% | 771 | 33% | 17 | 59% | 754 |  |
| Yes | 42% | 562 | 67% | 35 | 41% | 527 | <0.001 |
| **SHS visit in last year** |  |  |  |  |  |  |  |
| No | 36% | 478 | 12% | 6 | 37% | 472 |  |
| Yes | 64% | 855 | 88% | 46 | 63% | 809 | <0.001 |
| **STI tested in last year** |  |  |  |  |  |  |  |
| No | 30% | 405 | 10% | 5 | 31% | 400 |  |
| Yes | 70% | 928 | 90% | 47 | 69% | 881 | 0.001 |
| **STI test positivity since August 2022** |  |  |  |  |  |  |  |
| Never STI tested | 9% | 123 | 2% | 1 | 10% | 122 |  |
| Tested before August 2022 | 38% | 513 | 27% | 14 | 39% | 499 |  |
| All negative/unknown recent test results since August 2022 | 41% | 541 | 37% | 19 | 41% | 522 |  |

|  |  |  |  |  |  |  |  |  |  |  |
| --- | --- | --- | --- | --- | --- | --- | --- | --- | --- | --- |
| ≥1 recent positive test results since August 2022 | 12% | 156 |  | 35% | 18 |  | 11% | 138 |  | <0.001 |
| **Recreational drug use associated with chemsex in last year§** |  |  |  |  |  |  |  |  |  |  |
| No | 92% | 1222 |  | 79% | 41 |  | 92% | 1181 |  |  |
| Yes | 8% | 111 |  | 21% | 11 |  | 8% | 100 |  | 0.001 |
| **Number of recent male physical sex partners since August 2022** |  |  |  |  |  |  |  |  |  |  |
| None | 8% | 100 |  | 2% | 1 |  | 8% | 99 |  |  |
| 1 | 14% | 180 |  | 6% | 3 |  | 14% | 177 |  |  |
| 2 to 4 | 29% | 389 |  | 23% | 12 |  | 29% | 377 |  |  |
| 5 to 9 | 21% | 280 |  | 21% | 11 |  | 21% | 269 |  |  |
| ≥10 | 29% | 384 |  | 48% | 25 |  | 28% | 359 |  | 0.015 |
| **Met recent male physical sex partners in sex on premises venue, sex party, or PSE since Aug 2022** |  |  |  |  |  |  |  |  |  |  |
| No | 67% | 888 |  | 46% | 24 |  | 67% | 864 |  |  |
| Yes | 33% | 445 |  | 54% | 28 |  | 33% | 417 |  | 0.001 |
| **PrEP use since December 2021 (HIV negative/unknown status, n=1131)‡** |  |  |  |  |  |  |  |  |  |  |
| No | 47% | 534 |  | 17% | 6 |  | 48% | 528 |  |  |
| Yes | 53% | 597 |  | 83% | 30 |  | 52% | 567 |  | <0.001 |
| **Behaviour modification since May 2022 to avoid getting mpox¶** |  |  |  |  |  |  |  |  |  |  |
| No | 47% | 626 |  | 50% | 26 |  | 47% | 600 |  |  |
| Yes | 53% | 707 |  | 50% | 26 |  | 53% | 681 |  | 0.654 |
| *Participants with self-reported mpox test positivity. Those without mpox test positivity (i.e. no diagnosis history) include those without a known positive test (n=67) and those without a test history (n=1231). †Includes those identifying as bisexual, straight, or another way. ‡Excludes people living with HIV (n=202). § Includes crystal methamphetamine, mephedrone or gamma-hydroxybutyrate/gamma-butyrolactone. ¶Behaviour modification includes self-report of any of the following from May 2022: fewer sexual partners, reduced visits to sex on premises venues or PSE (i.e. cruising grounds), and avoiding: all sex, condomless anal sex, skin-to-skin contact, or visiting clubs or crowds. HAV=Hepatitis A virus. HBV=Hepatitis B virus. HPV=Human papilloma virus. SHS=Sexual health service. STI=Sexually transmitted infection. PSE=Public sex environment (i.e. cruising grounds). PrEP=HIV pre-exposure prophylaxis. Percentages may not total 100 due to rounding. |  |  |  |  |  |  |  |  |  |  |

**Appendix V.** Sociodemographic, clinical and behavioural characteristics of, and factors associated with self-reported mpox vaccination uptake among RiiSH-Mpox participants assumed eligible for mpox vaccination, November/December 2022.

|  |  |  |  |  |  |  |  |  |  |  |  |  |  |  |  |
| --- | --- | --- | --- | --- | --- | --- | --- | --- | --- | --- | --- | --- | --- | --- | --- |
|  | **% RiiSH-Mpox participants assumed vaccine eligible\*** |  | **% Mpox vaccinated†** |  |  |  |  |  |  |  |  |  |  |  |  |
| **Total** | **[column %]** | **n** | **[row %]** | **n** | **uOR** | **95% CI** |  |  | **p-value** |  | **aOR\*\*** | **95% CI** |  |  | **p-value** |
|  | 100% | 875 | 69% | 601 |  |  |  |  |  |  |  |  |  |  |  |
| **Sociodemographic characteristics** |  |  |  |  |  |  |  |  |  |  |  |  |  |  |  |
| Median age (IQR, range) | 45 (IQR: 36-54; range: 17-78) |  | 45 (IQR: 36-53; range: 18-74) |  |  |  |  |  |  |  |  |  |  |  |  |
| Mean age (standard deviation) | 44.9 (12.0) |  | 44.7 (11.7) |  |  |  |  |  |  |  |  |  |  |  |  |
| **Age-group‡** |  |  |  |  |  |  |  |  |  |  |  |  |  |  |  |
| 16-24 | 3% | 29 | 55% | 16 | 0.44 | 0.20 | - | 0.97 |  |  | 0.57 | 0.25 | - | 1.33 |  |
| 25-34 | 19% | 164 | 71% | 117 | 0.89 | 0.56 | - | 1.39 |  |  | 0.88 | 0.55 | - | 1.40 |  |
| 35-44 | 28% | 243 | 68% | 166 | 0.77 | 0.51 | - | 1.15 |  |  | 0.72 | 0.48 | - | 1.09 |  |
| 45-54 | 25% | 221 | 74% | 163 | 1.00 | (base) |  |  |  |  | 1.00 | (base) |  |  |  |
| ≥55 | 25% | 218 | 64% | 139 | 0.63 | 0.42 | - | 0.94 | 0.0915 |  | 0.76 | 0.50 | - | 1.18 | 0.4711 |
| **Gender identity and sex at birth** |  |  |  |  |  |  |  |  |  |  |  |  |  |  |  |
| Cisgender male | 99% | 862 | 69% | 594 | 1.00 | (base) |  |  |  |  | 1.00 | (base) |  |  |  |
| All other gender groups | 1% | 13 | 54% | 7 | 0.53 | 0.18 | - | 1.58 | 0.2603 |  | 0.75 | 0.24 | - | 2.38 |  |
| **Sexual orientation** |  |  |  |  |  |  |  |  |  |  |  |  |  |  |  |
| Gay/homosexual | 91% | 795 | 70% | 559 | 1.00 | (base) |  |  |  |  | 1.00 | (base) |  |  |  |
| Bisexual§ | 9% | 80 | 53% | 42 | 0.47 | 0.29 | - | 0.74 | 0.0015 |  | 0.49 | 0.30 | - | 0.79 | 0.0037 |
| **Ethnicity‡** |  |  |  |  |  |  |  |  |  |  |  |  |  |  |  |
| White | 92% | 802 | 69% | 552 | 1.00 | (base) |  |  |  |  | 1.00 | (base) |  |  |  |
| Other ethnicities | 8% | 73 | 67% | 49 | 0.92 | 0.55 | - | 1.54 | 0.7646 |  | 1.01 | 0.59 | - | 1.73 | 0.9749 |
| **Country of birth** |  |  |  |  |  |  |  |  |  |  |  |  |  |  |  |
| UK | 82% | 721 | 68% | 492 | 1.00 | (base) |  |  |  |  | 1.00 | (base) |  |  |  |
| Outside-UK | 18% | 154 | 71% | 109 | 1.13 | 0.77 | - | 1.65 | 0.081 |  | 1.03 | 0.68 | - | 1.54 | 0.8998 |
| **Nation of residence** |  |  |  |  |  |  |  |  |  |  |  |  |  |  |  |
| England | 87% | 760 | 69% | 528 | 1.00 | (base) |  |  |  |  | 1.00 | (base) |  |  |  |
| Scotland, Wales, or N Ireland | 13% | 115 | 63% | 73 | 0.76 | 0.51 | - | 1.15 | 0.2015 |  | 0.75 | 0.49 | - | 1.14 | 0.1821 |
| **Educational qualifications** |  |  |  |  |  |  |  |  |  |  |  |  |  |  |  |
| Below degree-level | 33% | 291 | 57% | 165 | 0.44 | 0.33 | - | 0.60 |  |  | 0.46 | 0.34 | - | 0.63 |  |
| Degree-level or higher | 67% | 584 | 75% | 436 | 1.00 | (base) |  |  | <0.001 |  | 1.00 | (base) |  |  | <0.001 |
| **Employment** |  |  |  |  |  |  |  |  |  |  |  |  |  |  |  |
| Not employed | 16% | 144 | 56% | 80 | 0.50 | 0.35 | - | 0.73 |  |  | 0.63 | 0.42 | - | 0.95 |  |
| Current employment | 84% | 731 | 71% | 521 | 1.00 | (base) |  |  | 0.0003 |  | 1.00 | (base) |  |  | 0.0286 |
| **Household composition** |  |  |  |  |  |  |  |  |  |  |  |  |  |  |  |
| Lives with others | 60% | 527 | 68% | 358 | 0.92 | 0.68 | - | 1.23 |  |  | 0.91 | 0.67 | - | 1.24 |  |
| Lives alone | 40% | 348 | 70% | 243 | 1.00 | (base) |  |  | 0.5535 |  | 1.00 | (base) |  |  | 0.5565 |
| **Relationship status** |  |  |  |  |  |  |  |  |  |  |  |  |  |  |  |
| Single | 45% | 397 | 71% | 281 | 1.20 | 0.90 | - | 1.60 |  |  | 1.24 | 0.92 | - | 1.68 |  |
| In a relationship | 55% | 478 | 67% | 320 | 1.00 | (base) |  |  | 0.2227 |  | 1.00 | (base) |  |  | 0.1571 |
| **Comfortable financial situation** |  |  |  |  |  |  |  |  |  |  |  |  |  |  |  |
| No | 48% | 420 | 66% | 278 | 0.80 | 0.60 | - | 1.07 |  |  | 1.00 | 0.74 | - | 1.35 |  |
| Yes (top two quartiles) | 52% | 455 | 71% | 323 | 1.00 | (base) |  |  | 0.1263 |  | 1.00 | (base) |  |  | 0.9896 |
| **Clinical and behavioural characteristics** |  |  |  |  |  |  |  |  |  |  |  |  |  |  |  |
| **HIV status** |  |  |  |  |  |  |  |  |  |  |  |  |  |  |  |
| Negative/unknown | 87% | 759 | 69% | 523 | 1.00 | (base) |  |  |  |  | 1.00 | (base) |  |  |  |
| Living with HIV | 13% | 116 | 67% | 78 | 0.93 | 0.61 | - | 1.41 | 0.7197 |  | 1.02 | 0.66 | - | 1.58 | 0.9269 |

| HAV, HBV, or HPV vaccination history¶ |  |  |  |  |  |  |  |  |  |  |  |  |  |  |  |
| --- | --- | --- | --- | --- | --- | --- | --- | --- | --- | --- | --- | --- | --- | --- | --- |
| No | 10% | 91 | 35% | 32 | 1.00 | (base) |  |  |  |  | 1.00 | (base) |  |  |  |
| Yes | 90% | 784 | 73% | 569 | 4.88 | 3.09 | - | 7.71 | <0.001 |  | 4.42 | 2.74 | - | 7.13 | <0.001 |
| Behaviour modification since May 2022 to avoid getting mpox# |  |  |  |  |  |  |  |  |  |  |  |  |  |  |  |
| No | 45% | 398 | 62% | 246 | 1.00 | (base) |  |  |  |  | 1.00 | (base) |  |  |  |
| Yes | 55% | 477 | 74% | 355 | 1.80 | 1.35 | - | 2.40 | 0.0001 |  | 1.63 | 1.21 | - | 2.20 | 0.0013 |

\*Mpox vaccine eligibility was defined as the report of any of the following since August 2022: $\geq 10$ physical male sex partners; meeting any physical male sex partners in a sex on premises venue, sex party, or PSE (i.e. cruising grounds); a positive STI test; or in the last year, report of: PrEP use (as a proxy for those at risk of mpox acquisition); or use of recreational drugs associated with chemsex (crystal methamphetamine, mephedrone or gamma-hydroxybutyrate/gamma-butyrolactone). †Participants with self-reported uptake of $\geq 1$ mpox vaccine dose(s). ‡Selected *a priori* for model inclusion. §Includes those identifying as bisexual, straight, or another way. ¶Participants self-reporting uptake of $\geq 1$ HAV, HBV, or HPV vaccine dose(s). #Behaviour modification includes self-report of any of the following from May 2022: fewer sexual partners, reduced visits to sex on premises venues or PSE (i.e. cruising grounds), and avoiding: all sex, condomless anal sex, skin-to-skin contact, or visiting clubs or crowds. \*\*Adjusted for age-group, ethnicity, sexual orientation, educational qualifications, employment. HAV=Hepatitis A virus. HBV=Hepatitis B virus. HPV=Human papilloma virus. SHS=Sexual health service. uOR=Unadjusted odds ratio. aOR=adjusted odds ratio. Percentages may not total 100 due to rounding.

**Appendix VI.**
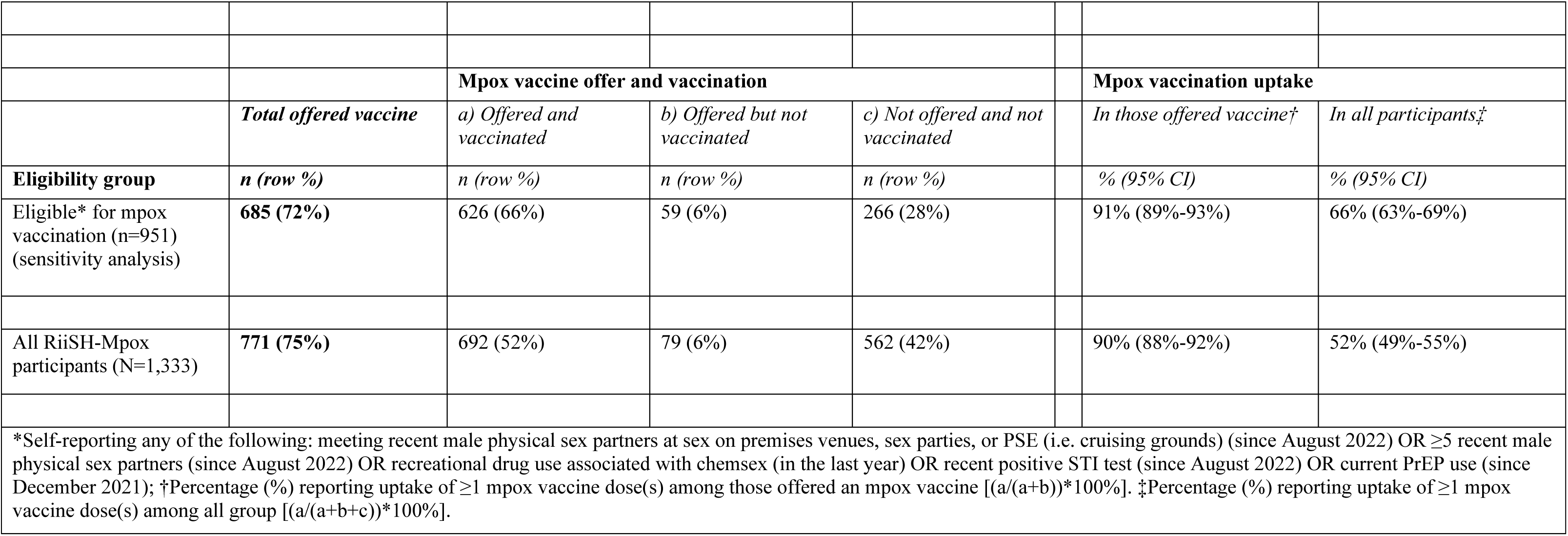
Mpox vaccine offer and uptake among RiiSH-Mpox participants considered eligible for mpox vaccination (sensitivity analysis, ≥5 recent male physical sex partners), November/December 2022.

|  |  |  |  |  |  |  |
| --- | --- | --- | --- | --- | --- | --- |
|  |  | **Mpox vaccine offer and vaccination** |  |  | **Mpox vaccination uptake** |  |
|  | ***Total offered vaccine*** | *a) Offered and vaccinated* | *b) Offered but not vaccinated* | *c) Not offered and not vaccinated* | *In those offered vaccine†* | *In all participants‡* |
| **Eligibility group** | ***n (row %)*** | ***n (row %)*** | ***n (row %)*** | ***n (row %)*** | ***% (95% CI)*** | ***% (95% CI)*** |
| Eligible* for mpox vaccination (n=951) (sensitivity analysis) | **685 (72%)** | 626 (66%) | 59 (6%) | 266 (28%) | 91% (89%-93%) | 66% (63%-69%) |
| All RiiSH-Mpox participants (N=1,333) | **771 (75%)** | 692 (52%) | 79 (6%) | 562 (42%) | 90% (88%-92%) | 52% (49%-55%) |
| *Self-reporting any of the following: meeting recent male physical sex partners at sex on premises venues, sex parties, or PSE (i.e. cruising grounds) (since August 2022) OR ≥5 recent male physical sex partners (since August 2022) OR recreational drug use associated with chemsex (in the last year) OR recent positive STI test (since August 2022) OR current PrEP use (since December 2021); †Percentage (%) reporting uptake of ≥1 mpox vaccine dose(s) among those offered an mpox vaccine [(a/(a+b))*100%]. ‡Percentage (%) reporting uptake of ≥1 mpox vaccine dose(s) among all group [(a/(a+b+c))*100%]. |  |  |  |  |  |  |

## Notes

### Competing Interest Statement

The authors have declared no competing interest.

## References

1. Ghebreyesus TA, World Health O. Why the monkeypox outbreak constitutes a public health emergency of international concern. BMJ. 2022 Aug 9;378:o1978.

2. Centers for Disease Control and Prevention. 2022 Global Map & Case Count. 2023 [cited 2023 24 March]; Available from: https://www.cdc.gov/poxvirus/mpox/response/2022/world-map.html

3. The UK Health Security Agency (UKHSA). UK strategy for mpox control, 2022 to 2023. 2022 [cited 2023 24 March]; Available from: https://www.gov.uk/government/publications/mpox-monkeypox-control-uk-strategy-2022-to-2023/uk-strategy-for-mpox-control-2022-to-2023

4. Inigo Martinez J, Gil Montalban E, Jimenez Bueno S, Martin Martinez F, Nieto Julia A, Sanchez Diaz J, et al. Monkeypox outbreak predominantly affecting men who have sex with men, Madrid, Spain, 26 April to 16 June 2022. Euro Surveill. 2022 Jul;27(27).

5. Vivancos R, Anderson C, Blomquist P, Balasegaram S, Bell A, Bishop L, et al. Community transmission of monkeypox in the United Kingdom, April to May 2022. Euro Surveill. 2022 Jun;27(22):2200422.

6. The UK Health Security Agency (UKHSA). Mpox (monkeypox) outbreak vaccination strategy. 2022 [cited 2023 24 March]; Available from: https://www.gov.uk/guidance/monkeypox-outbreak-vaccination-strategy

7. The UK Health Security Agency (UKHSA). Mpox and sexual health: outreach and engagement fund. 2022 [cited 2023 24 April]; Available from: https://www.gov.uk/government/publications/mpox-and-sexual-health-outreach-and-engagement-fund

8. Terrence Higgins Trust. Mpox (monkeypox) in the UK. 2023 [cited 2023 24 April]; Available from: https://www.tht.org.uk/hiv-and-sexual-health/sexual-health/mpox-monkeypox-uk

9. The Love Tank. 2023 [cited 2023 24 April]; Available from: http://thelovetank.info/

10. The UK Health Security Agency (UKHSA). Investigation into monkeypox outbreak in England: technical briefing 8. 2022 [cited 2023 24 March]; Available from: https://www.gov.uk/government/publications/monkeypox-outbreak-technical-briefings/investigation-into-monkeypox-outbreak-in-england-technical-briefing-8

11. NHS England. Vaccinations for mpox: MVA vaccinations 4 May 2023. 2023 [cited 2023 10 May]; Available from: https://www.england.nhs.uk/statistics/statistical-work-areas/vaccinations-for-mpox/

12. The UK Health Security Agency (UKHSA). Mpox (monkeypox) outbreak: epidemiological overview, 4 May 2023. 2023 [cited 2023 10 May]; Available from: https://www.gov.uk/government/publications/monkeypox-outbreak-epidemiological-overview

13. The UK Health Security Agency (UKHSA). Smallpox and monkeypox:the green book, chapter 29. 2022 [cited 2023 15 February]; Available from: https://assets.publishing.service.gov.uk/government/uploads/system/uploads/attachment_data/file/1106454/Green-Book-chapter-29_Smallpox-and-monkeypox_26September2022.pdf

14. The UK Health Security Agency (UKHSA). People still eligible for mpox vaccine urged to come forward. 2023 [cited 2023 24 March]; Available from: https://www.gov.uk/government/news/people-still-eligible-for-mpox-vaccine-urged-to-come-forward

15. The UK Health Security Agency (UKHSA). Monkeypox vaccines to be piloted in smaller but equally effective doses. 2022 [cited; Available from: https://www.gov.uk/government/news/monkeypox-vaccines-to-be-piloted-in-smaller-but-equally-effective-doses

16. Bertran M, Andrews N, Davison C, Dugbazah B, Boateng J, Lunt R, et al. Effectiveness of one dose of MVA-BN smallpox vaccine against mpox in England using the case-coverage method: an observational study. Lancet Infect Dis. 2023 Mar 13.

17. Wolff Sagy Y, Zucker R, Hammerman A, Markovits H, Arieh NG, Abu Ahmad W, et al. Real-world effectiveness of a single dose of mpox vaccine in males. Nat Med. 2023 Mar;29(3):748–52.

18. Brown JR, Reid D, Howarth AR, Mohammed H, Saunders J, Pulford CV, et al. Changes in STI and HIV testing and testing need among men who have sex with men during the UK’s COVID-19 pandemic response. Sex Transm Infect. 2022 Jul 21.

19. Howarth AR, Saunders J, Reid D, Kelly I, Wayal S, Weatherburn P, et al. ’Stay at home …’: exploring the impact of the COVID-19 public health response on sexual behaviour and health service use among men who have sex with men: findings from a large online survey in the UK. Sex Transm Infect. 2022 Aug;98(5):346–52.

20. Curran KG, Eberly K, Russell OO, Snyder RE, Phillips EK, Tang EC, et al. HIV and Sexually Transmitted Infections Among Persons with Monkeypox - Eight US Jurisdictions, May 17-July 22, 2022. Mmwr-Morbidity and Mortality Weekly Report. 2022 Sep 9;71(36):1141–7.

21. Tarin-Vicente EJ, Alemany A, Agud-Dios M, Ubals M, Suner C, Anton A, et al. Clinical presentation and virological assessment of confirmed human monkeypox virus cases in Spain: a prospective observational cohort study. Lancet. 2022 Aug 27;400(10353):661–9.

22. Gilbert M, Ablona A, Chang HJ, Grennan T, Irvine MA, Sarai Racey C, et al. Uptake of Mpox vaccination among transgender people and gay, bisexual and other men who have sex with men among sexually-transmitted infection clinic clients in Vancouver, British Columbia. Vaccine. 2023 Apr 6;41(15):2485–94.

23. Nadarzynski T, Nutland W, Samba P, Bayley J, Witzel TC. The Impact of First UK-Wide Lockdown (March-June 2020) on Sexual Behaviors in Men and Gender Diverse People Who Have Sex with Men During the COVID-19 Pandemic: A Cross-Sectional Survey. Arch Sex Behav. 2023 Feb;52(2):617–27.

24. Thorley K, Charles H, Greig DR, Prochazka M, Mason LCE, Baker KS, et al. Emergence of extensively drug-resistant and multidrug-resistant Shigella flexneri serotype 2a associated with sexual transmission among gay, bisexual, and other men who have sex with men, in England: a descriptive epidemiological study. Lancet Infect Dis. 2023 Jan 30.

25. Ogaz D, Allen H, Reid D, Brown JRG, Howarth AR, Pulford CV, et al. COVID-19 infection and vaccination uptake in men and gender-diverse people who have sex with men in the UK: analyses of a large, online community cross-sectional survey (RiiSH-COVID) undertaken November-December 2021. BMC Public Health. 2023 May 5;23(1):829.

26. Curtis T, Bennett K, Mcdonagh L, Field N, Mercer C. P532 The sexual behaviour and health of heterosexual-identifying men who have sex with men: a systematic review. Sexually Transmitted Infections. 2019;95(Suppl 1):A242-A.

27. Desai S, Burns F, Schembri G, Williams D, Sullivan A, McOwan A, et al. Sexual behaviours and sexually transmitted infection outcomes in a cohort of HIV-negative men who have sex with men attending sexual health clinics in England. Int J STD AIDS. 2018 Dec;29(14):1407–16.

28. Mitchell HD, Desai S, Mohammed H, Ong KJ, Furegato M, Hall V, et al. Preparing for PrEP: estimating the size of the population eligible for HIV pre-exposure prophylaxis among men who have sex with men in England. Sex Transm Infect. 2019 Nov;95(7):484–7.

29. O’Halloran C, Owen G, Croxford S, Sims LB, Gill ON, Nutland W, et al. Current experiences of accessing and using HIV pre-exposure prophylaxis (PrEP) in the United Kingdom: a cross-sectional online survey, May to July 2019. Euro Surveill. 2019 Nov;24(48):1900693.

30. Ogaz D, Logan L, Curtis TJ, McDonagh L, Guerra L, Bradshaw D, et al. PrEP use and unmet PrEP-need among men who have sex with men in London prior to the implementation of a national PrEP programme, a cross-sectional study from June to August 2019. BMC Public Health. 2022 Jun 3;22(1):1105.

31. Wise J. Monkeypox: UK to run out of vaccine doses by next week. BMJ. 2022 Aug 18;378:o2053.

32. Heskin J, Dickinson M, Brown N, Girometti N, Feeney M, Hardie J, et al. Rapid reconfiguration of sexual health services in response to UK autochthonous transmission of mpox (monkeypox). Sex Transm Infect. 2023 Mar;99(2):81–4.

33. Jamard S, Handala L, Faussat C, Vincent N, Stefic K, Gaudy-Graffin C, et al. Resurgence of symptomatic Mpox among vaccinated patients: First clues from a new-onset local cluster. Infect Dis Now. 2023 Apr 28;53(4):104714.

34. Department of Health and Social Care. JCVI statement on vaccine dose prioritisation in response to the monkeypox outbreak. 2022 [cited 2023 01 April]; Available from: https://www.gov.uk/government/publications/monkeypox-outbreak-jcvi-statement-on-vaccine-dose-prioritisation-september-2022/jcvi-statement-on-vaccine-dose-prioritisation-in-response-to-the-monkeypox-outbreak#updated-advice-september-2022

35. Paparini S, Whitacre R, Smuk M, Thornhill J, Mwendera C, Strachan S, et al. Public understanding and awareness of and response to monkeypox virus outbreak: A cross-sectional survey of the most affected communities in the United Kingdom during the 2022 public health emergency. HIV Med. 2022 Nov 16.

36. Dukers-Muijrers N, Evers Y, Widdershoven V, Davidovich U, Adam PCG, Op de Coul ELM, et al. Mpox vaccination willingness, determinants, and communication needs in gay, bisexual, and other men who have sex with men, in the context of limited vaccine availability in the Netherlands (Dutch Mpox-survey). Front Public Health. 2022;10:1058807.

37. MacGibbon J, Cornelisse V, Smith AKJ, Broady TR, Hammoud MA, Bavinton BR, et al. Mpox (monkeypox) knowledge, concern, willingness to change behaviour, and seek vaccination: Results of a national cross-sectional survey. medRxiv. 2023:2022.12.01.22282999.

38. Mercer CH, Prah P, Field N, Tanton C, Macdowall W, Clifton S, et al. The health and well-being of men who have sex with men (MSM) in Britain: Evidence from the third National Survey of Sexual Attitudes and Lifestyles (Natsal-3). BMC Public Health. 2016 Jul 7;16(1):525.

39. Queer Health. Everything we know about MPOX (Monkeypox) so far. 2023 [cited 2023 20 April]; Available from: https://www.queerhealth.info/projects/monkeypox

40. Logie CH. What can we learn from HIV, COVID-19 and mpox stigma to guide stigma-informed pandemic preparedness? J Int AIDS Soc. 2022 Dec;25(12):e26042.

